# Natural-language retrieval with multimodal embeddings identifies candidate developmental behaviors in caregiver-child recordings

**DOI:** 10.64898/2026.08.09.26360057

**Authors:** Benson Mwangi, Mon-Ju Wu, Rosleen Mansour, Gabriel Anzueto, Antonio F. Pagán

## Abstract

**Background:** Naturalistic audiovisual recordings of caregiver-child interactions contain rich developmental signals. However, extracting interpretable clinical measures requires resource-intensive manual coding. To address this bottleneck, we evaluated natural-language queries for retrieving specific behavioral moments from these recordings, applying multimodal embeddings as an automated evidence-selection layer.

**Methods:** We compared three embedding models (Jina Embeddings v5 Omni, LanguageBind, and Wave7B) for natural-language retrieval directly from audio and video streams, bypassing transcript text. We assessed performance across 27 behavioral targets in 277 caregiver-child recordings (14, 24, and 36 months of age) from the Early Head Start Talkbank corpus, yielding 7,479 recording-target queries.

**Results:** Jina Embeddings v5 Omni achieved the highest top-10 retrieval success (text-to-audio 38.3%; text-to-video 36.4%), ahead of LanguageBind (37.0%; 34.5%) and Wave7B (36.1%; 35.0%). Across models, retrieval was substantially more successful for common targets than for rare vocal and gestural behaviors, such as pointing and babbling. By analyzing the spoken words within the retrieved audio clips, we found that Jina accurately ranked the children by their relative vocabulary size at each age (Spearman ⍴ = 0.68, 0.82, and 0.90 at 14, 24, and 36 months). However, the model severely underestimated the total number of unique words each child used throughout the full session.

**Conclusion:** Multimodal embeddings can successfully pinpoint important developmental behaviors and speech patterns within lengthy caregiver-child recordings. However, these systems still struggle to locate rare events. Additionally, while they can accurately rank children by relative vocabulary size, they fail to measure a child’s complete vocabulary. We conclude that these models are currently best suited for automated evidence-selection to prioritize relevant segments for expert interpretation rather than acting as an independent replacement for manual behavioral coding or language assessment. Improving the detection of infrequent behaviors and validating these models across external datasets are essential next steps before real-world clinical deployment.

## Introduction

Developmental disabilities affect approximately one in six children in the United States (Zablotsky et al. 2019). Although standardized screening tools offer valuable structured clinical indicators, timely identification still depends heavily on directly observing naturalistic interactions between caregivers and children (Lipkin et al. 2020; Gardner 2000). These everyday exchanges reveal essential behavioral markers that are critical for developmental surveillance. For example, responsive caregiving and sustained joint attention are strong prospective predictors of early expressive language milestones (Tomasello and Farrar 1986). Similarly, the quality of a caregiver’s linguistic input directly scaffolds a child’s vocabulary acquisition (Golinkoff et al. 2015). Early gestures like pointing reliably forecast future lexical and syntactic development (Choi et al. 2021; Rowe and Goldin-Meadow 2009). Furthermore, reciprocal interactions that contingently reinforce early babbling actively shape mature vocal behavior (Goldstein et al. 2003). Conversational turn-taking is longitudinally tied to vocabulary growth, and coaching parents to enhance these dyadic interactions has been shown to reliably increase a child’s speech complexity (Donnelly and Kidd 2021).

Home-based audiovisual recordings extend the reach of direct observation by preserving these behaviors for subsequent review. However, extracting interpretable data from these sessions remains prohibitively resource-intensive. Speech must be manually transcribed, while gestures, affective displays, and interaction events require rigorous coding by trained personnel (Miller et al. 2011; Pezold et al. 2020; Wittenburg et al. 2006). This heavy reliance on human expertise severely limits scalability across broad research cohorts and early childhood services. Existing automated methods address segments of this challenge, but their outputs are typically constrained to fixed taxonomies or aggregate statistics. Audio-based systems can estimate speaker-specific vocal activity and tally phoneme, syllable, and word counts from daylong child-centered recordings. While useful, their performance remains sensitive to diarization errors, overlapping speech, and complex acoustic environments (Räsänen et al. 2021; Cristia et al. 2021; Gilkerson et al. 2017). Crucially, these systems provide overarching summaries rather than natural-language access to specific behavioral events. Similarly, video-based systems can quantify emotion or spatial dynamics (Egger et al. 2018; Isaev et al. 2024; Weng et al. 2025), and multimodal models can predict predefined infant-caregiver engagement states (Withanage Don et al. 2024) Ultimately, session-level scores and static classifications fail to isolate the contextual triggers, event sequences, and reciprocal responses necessary for rigorous clinical interpretation.

Cross-modal retrieval addresses this localization bottleneck by using natural-language descriptions to search audio and video directly. These systems map text queries and temporally segmented audiovisual content into an aligned vector embedding space, ranking the segments based on their semantic similarity to the queried behavior (Girdhar et al. 2023; Zhu et al. 2023). Unlike fixed classifiers, cross-modal retrieval allows a single recording to be queried for multiple behavioral concepts without requiring target-specific model architectures. Crucially, the retrieved behavior remains linked to its original audiovisual context, establishing a highly scalable evidence-selection layer for developmental measurement.

Applying this technology to naturalistic interactions introduces technical challenges that are rarely captured by conventional retrieval benchmarks. Target clinical signals such as infant vocalizations are often sparse, brief, acoustically ambiguous or heavily overlapped with caregiver speech. Home recording conditions also introduce high variance in background noise, illumination and camera occlusion. Developmentally informative behaviors like babbling or pointing are fleeting and underrepresented in general-purpose multimodal pretraining datasets, collectively generating a severe domain shift. State-of-the-art performance on standard benchmarks therefore does not guarantee reliable clinical measurement.

Furthermore, the retrieval target itself exhibits profound developmental nonstationarity. At 14 months, communication relies heavily on prelinguistic vocalizations, gestures and emerging single words. By 24 months, vocabulary expands and early word combinations appear. By 36 months, multiword speech fundamentally alters the available audiovisual evidence (Brown 2009; Fenson and Others 2007). Longitudinal evaluation is therefore essential to determine whether retrieval reliability persists as a child’’s communicative capacity matures. Finally, locating relevant audiovisual moments is a necessary but insufficient step for assessment. Validating the clinical utility of the system requires testing whether it preserves established language metrics such as vocabulary diversity, mean length of utterance and single-word proportion to distinguish basic evidence localization from accurate quantitative measurement (Brown 1973; Watkins et al. 1995).

Here, we evaluate phrase-based cross-modal retrieval as an automated evidence-selection layer for developmental language measurement. We compared three open-weight multimodal embedding models; LanguageBind (Zhu et al. 2023), Wave7B (Tang et al. 2025), and Jina Embeddings v5 Omni (Hönicke et al. 2026), using short natural-language descriptions to search temporally segmented content directly from audio and video streams. Assessing 27 behavioral targets in 277 caregiver-child recordings from the Early Head Start corpus spanning 14, 24, and 36 months of age, we quantified retrieval performance across models, developmental stages, and behavioral classes (MacWhinney 2007; Pan, B. A., Ayoub C., Snow, C. E. 2008; Pan et al. 2005). We subsequently tested whether language measures derived from the retrieved segments successfully preserved the relative differences and absolute scales of baseline human-coded metrics. Through this framework, we isolate the specific multimodal capabilities that require refinement prior to clinical deployment.

### 2.1 Study Design and Overview

This retrospective computational study evaluated phrase-based cross-modal retrieval from archived mother-child recordings at 14, 24, and 36 months of age. We addressed two primary objectives. First, we evaluated whether short natural-language queries could retrieve temporally localized, behaviorally relevant audiovisual segments. We compared three multimodal embedding models (LanguageBind, Wave7B, and Jina Embeddings v5 Omni) under identical zero-shot conditions, utilizing longitudinal models to assess performance across developmental waves, behavioral classes, and target modalities. Second, we assessed whether child speech segments localized through retrieval preserved established developmental language metrics such as vocabulary diversity, mean length of utterance, and single-word proportion. By comparing retrieval-derived metrics against complete-session reference values calculated from human transcripts, we distinguished basic evidence localization from true quantitative measurement.

### 2.2 Data Source and Participants

We utilized data from the National Evaluation of Early Head Start corpus (Harvard-Brattleboro, Vermont site), archived within the TalkBank Child Language Data Exchange System (CHILDES) (MacWhinney 2007; Pan et al. 2005). The source cohort comprised predominantly European American, English-speaking, low-income families from rural New England (Pan et al. 2005). Individual-level demographics and program assignments were unavailable for our specific analytical sample. The initial archive contained 279 wave-specific transcripts. After excluding two 14-month files lacking processed recording directories, the final analytic sample comprised 277 recordings (105 at 14 months, 94 at 24 months, and 78 at 36 months) linked across 121 distinct participant identifiers. We evaluated each recording against 27 prespecified targets, yielding 7,479 recording-target queries.

The observational protocol featured a 10-minute three-bags semi-structured play interaction (Cabrera et al. 2007; Rowe et al. 2017). Because some recordings included supplementary tasks such as frustration, teaching, or free-play exercises, recording durations and behavioral compositions varied longitudinally (Pan, B. A., Ayoub C., Snow, C. E. 2008; Pan et al. 2005). Recordings were transcribed using standard CHILDES CHAT conventions (MacWhinney 2017), capturing timestamped verbal utterances, nonverbal communicative actions, activity contexts, and paralinguistic events like crying, babbling, and pointing. The source corpus preparation included secondary transcriber verification, with pointing annotations demonstrating 84% inter-rater agreement in a 10% random subsample (Pan et al. 2005; MacWhinney 2019). We utilized these archived annotations to derive ground-truth chunk-level reference labels and complete-session developmental metrics. We parsed the archived CHAT files directly rather than through childes-db because the media links, sparse annotation tiers, reformulations, and paralinguistic material required for chunk-level reference construction are not fully represented in that database interface (Sanchez et al. 2019).

### 2.3 Recording Preprocessing and Temporal Segmentation

We partitioned each recording into consecutive, non-overlapping 20-second chunks and retained any residual terminal intervals. This process yielded 20,195 chunks totaling 111.45 hours (Table 2). For every chunk, we used the FFmpeg software library (Lei et al. 2013) to extract synchronized video-only (MP4) and audio-only (16-bit PCM WAV, 16 kHz) streams to support modality-specific and combined processing. We parsed the transcripts using the PyLangAcq library (Lee et al. 2016) and assigned each timestamped utterance to the chunk containing its onset. Using prespecified rules, we converted these archived annotations into 27 binary, multilabel reference tags that captured speaker identities, questioning, paralinguistic behaviors, and activity contexts (Table 1). We then designated the targets a priori as AudioPrimary or VideoPrimary based on the expected optimal retrieval modality.

**Table 1.**
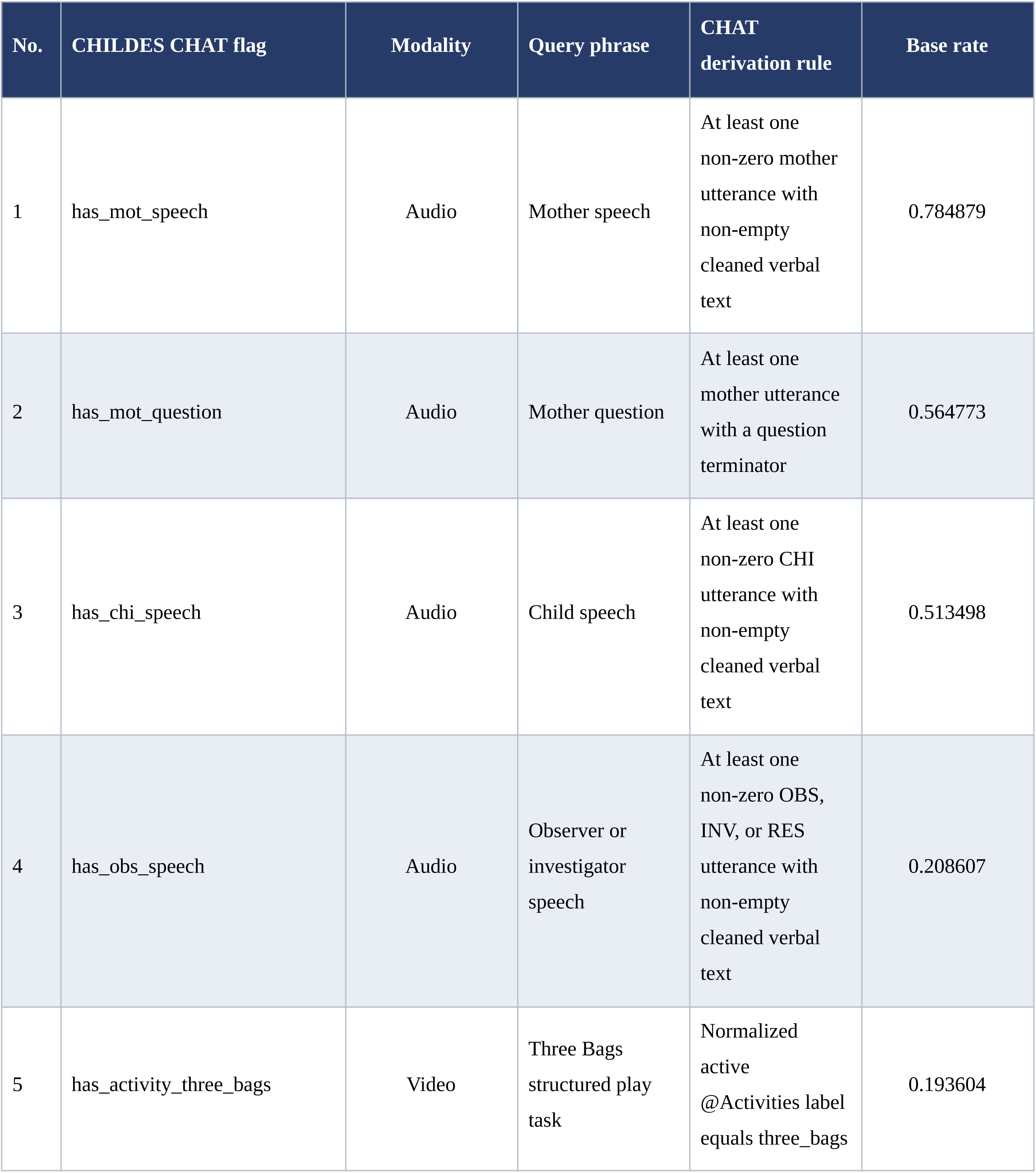
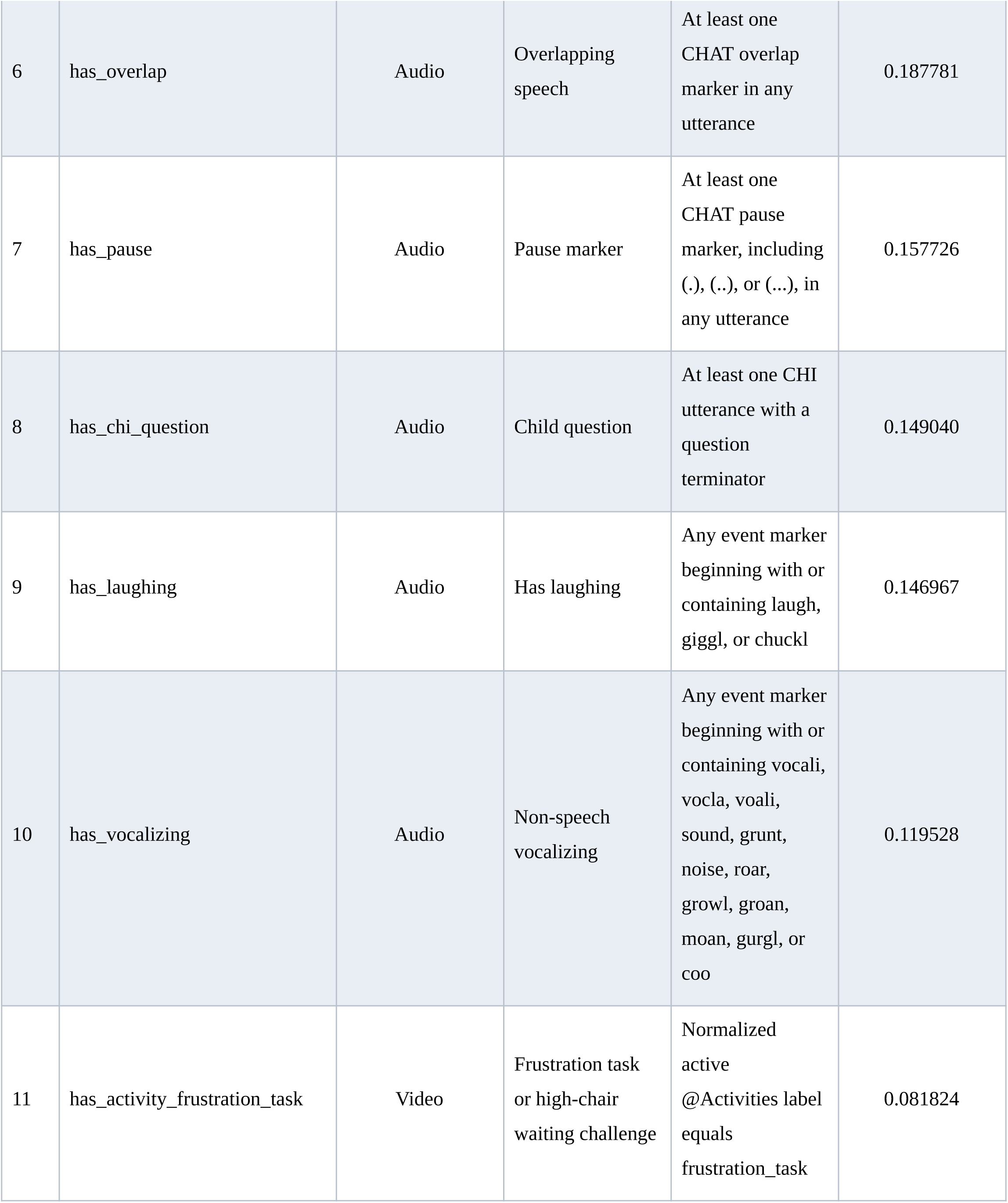
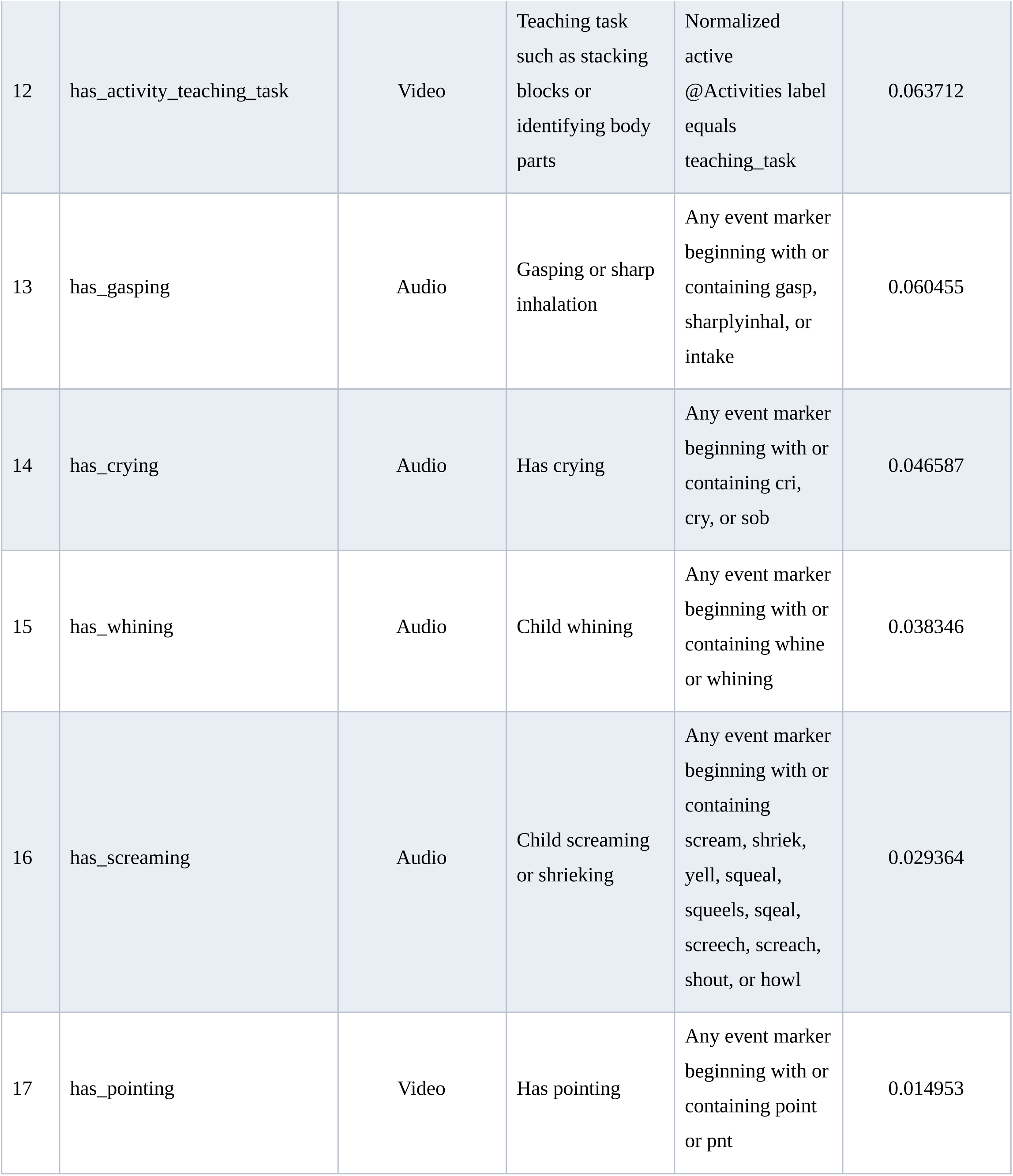
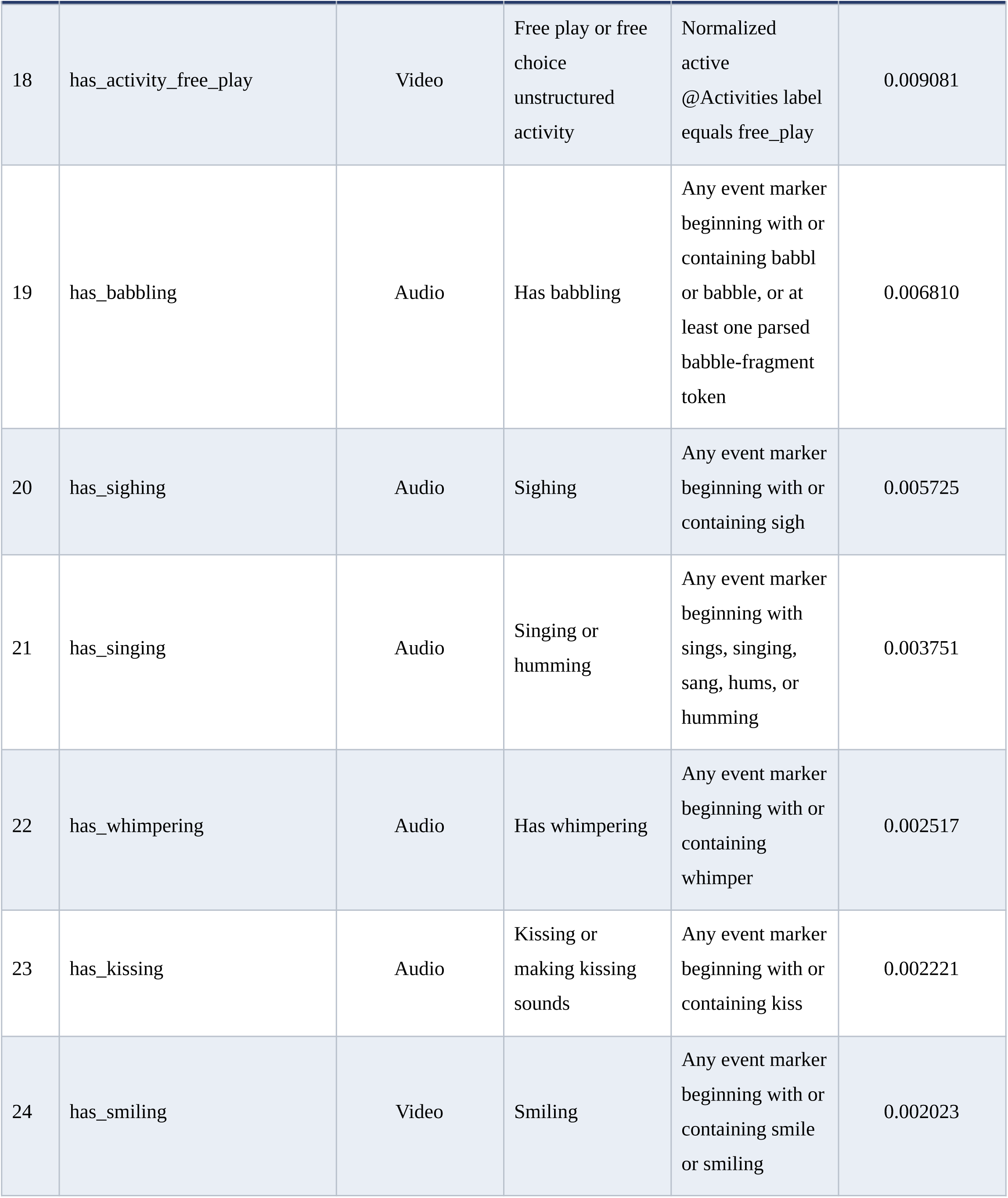
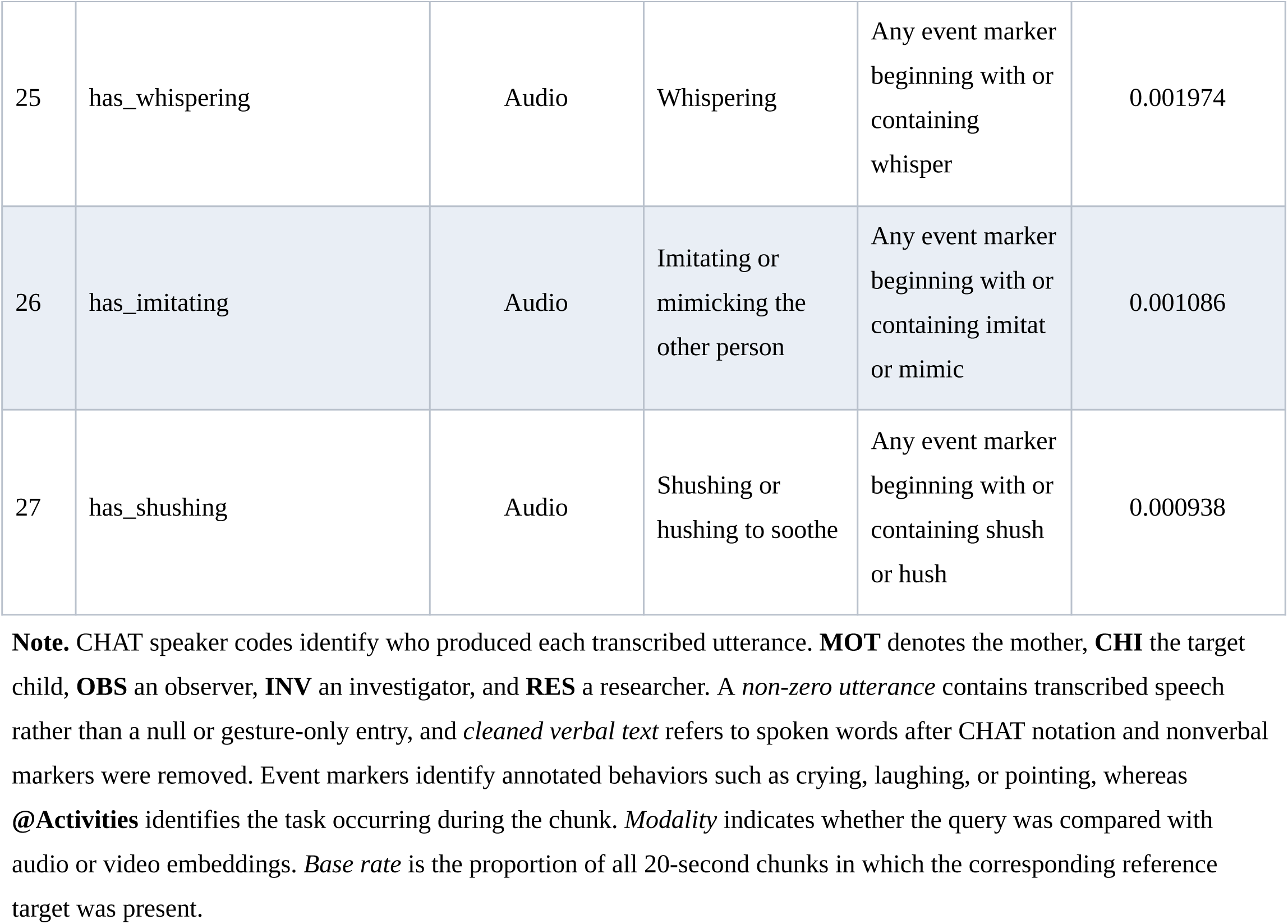
Behavioral target taxonomy, query phrases, CHAT derivation rules, and combined-cohort base rates.

**Table 2.**
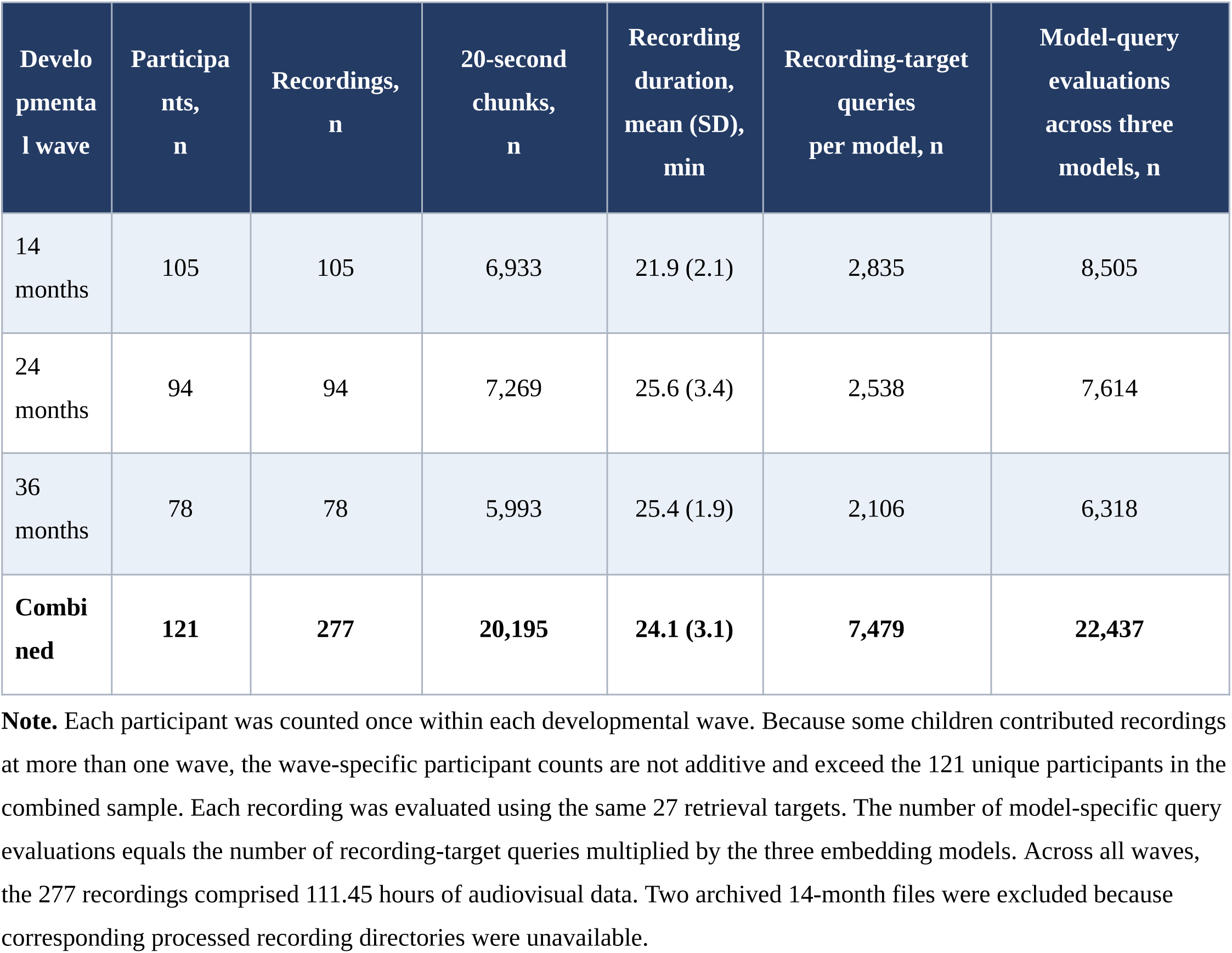
Analytic sample, recording duration, segmentation, and retrieval evaluation volume by developmental wave.

| Developmental wave | Participants, n | Recordings, n | 20-second chunks, n | Recording duration, mean (SD), min | Recording-target queries per model, n | Model-query evaluations across three models, n |
| --- | --- | --- | --- | --- | --- | --- |
| 14 months | 105 | 105 | 6,933 | 21.9 (2.1) | 2,835 | 8,505 |
| 24 months | 94 | 94 | 7,269 | 25.6 (3.4) | 2,538 | 7,614 |
| 36 months | 78 | 78 | 5,993 | 25.4 (1.9) | 2,106 | 6,318 |
| **Combined** | **121** | **277** | **20,195** | **24.1 (3.1)** | **7,479** | **22,437** |

**Note.** Each participant was counted once within each developmental wave. Because some children contributed recordings at more than one wave, the wave-specific participant counts are not additive and exceed the 121 unique participants in the combined sample. Each recording was evaluated using the same 27 retrieval targets. The number of model-specific query evaluations equals the number of recording-target queries multiplied by the three embedding models. Across all waves, the 277 recordings comprised 111.45 hours of audiovisual data. Two archived 14-month files were excluded because corresponding processed recording directories were unavailable.

### 2.4 Audiovisual Indexing and Retrieval

LanguageBind (Zhu et al. 2023), Wave7B (Tang et al. 2025) and Jina Embeddings v5 Omni (Hönicke et al. 2026) independently processed each 20-second chunk. We indexed the recordings by developmental wave and stored the media in MinIO object storage (Gadban and Kunkel 2021). The corresponding audio and video embeddings were stored in a Qdrant vector database (Ockerman et al. 2025) and linked through shared chunk identifiers. Because each model maps text, audio and video into a shared embedding space, we directly compared the natural-language query text vectors against chunk-level audio vectors for AudioPrimary targets or video vectors for VideoPrimary targets. All embeddings were strictly isolated by model. During retrieval, Qdrant ranked chunks from the same recording based on their cosine similarity (Plale et al. 2025; Singh et al. 2023) to the query embedding and returned the top 10 unique candidates. Crucially, to prevent data leakage (Passos et al. 2019; Mwangi et al. 2014), transcript text was entirely excluded from the retrieval channel. Human-coded reference tags were accessed exclusively post-retrieval for performance evaluation (Fig. 1).

**Fig. 1.**
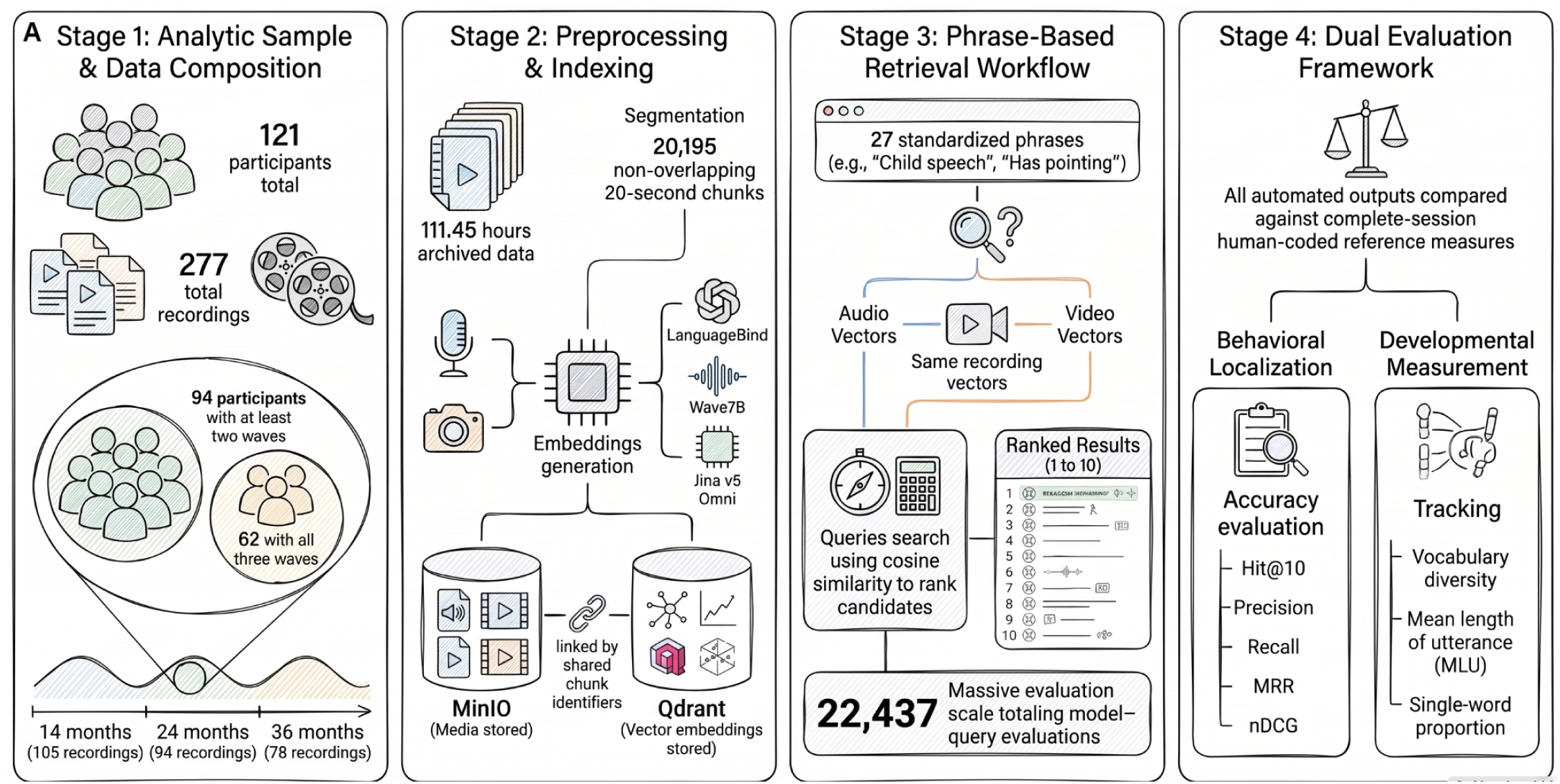
Study design and phrase-based cross-modal retrieval workflow. The analytic sample comprised 277 archived caregiver-child recordings from 121 participants, including 105 recordings at 14 months, 94 at 24 months, and 78 at 36 months. Ninety-four participants contributed recordings at two or more developmental waves, including 62 represented at all three waves. Recordings totaling 111.45 hours were partitioned into 20,195 non-overlapping 20-second chunks. Aligned media files were stored in MinIO, while LanguageBind, Wave7B, and Jina Embeddings v5 Omni generated model-specific audio and video embeddings that were stored in Qdrant and linked to the source media through shared chunk identifiers. For each recording, 27 standardized natural-language phrases were encoded and compared with audio or video vectors from the same recording using cosine similarity, yielding ranked candidate chunks. The evaluation comprised 7,479 recording-target queries per model and 22,437 model-query evaluations overall. Behavioral localization was assessed against chunk-level human-coded target labels, whereas retrieval-derived vocabulary diversity, mean length of utterance, and single-word proportion were compared with complete-session transcript-derived reference measures. The figure is schematic and does not display empirical performance results.

### 2.5 Multimodal Embedding Models

We selected three multimodal embedding models that represent distinct cross-modal alignment strategies. LanguageBind contrastively aligns modality-specific encoders with a language representation (Zhu et al. 2023). Wave7B derives prompt-aware representations through a multimodal language-model architecture (Tang et al. 2025). Finally, Jina Embeddings v5 Omni bridges pretrained perceptual encoders to a text backbone through lightweight projections (Hönicke et al. 2026). We evaluated all models using a zero-shot approach, applying them without any fine-tuning or adaptation to the Early Head Start corpus. Checkpoint identifiers, inference configurations and hardware environments are detailed in Supplementary Table S3.

### 2.6 Retrieval Outcomes

We evaluated retrieval performance independently for each queried target. Our primary endpoint was Hit@10, which equaled 1 if the ten highest-ranked chunks contained at least one target-positive match and 0 otherwise. We prioritized Hit@10 to establish the baseline success of locating any usable candidate evidence for human review. We subsequently applied secondary metrics to capture distinct dimensions of retrieval quality (Manning 2008). Precision@10 evaluated the signal-to-noise ratio by quantifying the proportion of retrieved chunks that contained the target. Recall@10 measured evidence completeness by calculating the fraction of all available target-positive chunks successfully retrieved. To quantify how rapidly a human reviewer would encounter the first relevant result, mean reciprocal rank (MRR) captured the reciprocal position of the initial positive chunk, scoring 0 if none were retrieved. To assess overall ranking utility when multiple relevant segments existed within a recording, we utilized normalized discounted cumulative gain at 10 (nDCG@10). This metric incorporated the positions of all positive chunks while heavily weighting earlier matches (Järvelin and Kekäläinen 2002). We queried all 27 targets in every recording regardless of their underlying behavioral presence. We classified queries as target-present if the recording contained at least one reference-positive chunk and target-absent otherwise. Target-absent queries inherently could not produce a retrieval hit. Consequently, while direct cross-model comparisons utilized the complete query panel, our longitudinal analyses isolated target-present queries to assess performance strictly when relevant evidence existed within the recording. We defined chunk-level target prevalence as the proportion of chunks bearing a positive reference tag for the corresponding target.

### 2.7 Reconstruction and Validation of Developmental Language Measures

For each recording and model, we linked the ten chunks retrieved using the standardized child-speech query to their corresponding human-transcribed child utterances. The retrieval system localized candidate segments but did not autonomously transcribe audio or compute language metrics. We calculated retrieval-derived measures strictly from utterances within the selected chunks and contrasted these against complete-session reference values derived from all eligible child utterances in the source recording. We defined child vocabulary diversity (CVD) as the count of unique lowercased word types across included utterances (Watkins et al. 1995). We calculated mean length of utterance (MLU) in words by dividing total child word tokens by the number of non-zero verbal utterances (Brown 1973). Single-word proportion (SWP) represented the fraction of verbal utterances containing exactly one word (Fenson and Others 2007). We considered MLU and SWP undefined in the absence of eligible verbal utterances. We applied identical definitions to the retrieved subsets and complete-session references. Because vocabulary diversity, mean length of utterance and single-word proportion are properties of the speech stream, our primary language validation used text-to-audio retrieval of child-speech queries and did not pool text-to-video rankings.

### 2.8 Statistical Analysis

#### Cross-model retrieval comparisons

For descriptive summaries, we averaged each model’s retrieval metrics across the common set of 7,479 recording-target queries to allow every query to contribute equally. To account for target clustering within recordings and repeated longitudinal observations of the same child, we conducted our statistical comparisons at the participant level. Within each developmental wave, we first averaged each retrieval metric across the 27 targets in a recording to yield one value per participant and model. For the combined analysis, we then averaged these recording-level values across all available developmental waves for each participant, producing a single summary value per participant, model, and retrieval metric.

We evaluated pairwise model differences using two-sided Wilcoxon signed-rank tests on the participant-level values. Participants presenting a paired difference of zero were excluded from the signed-rank calculations, and we reported the resulting number of non-zero pairs. We applied rank-biserial correlations to quantify the magnitude and direction of each paired difference. Next, we estimated mean paired differences alongside percentile 95% confidence intervals utilizing 2,000 participant-level bootstrap resamples.

We applied multiplicity adjustments across the 15 aggregate comparisons arising from five retrieval metrics and three model pairs. The Benjamini-Hochberg procedure (Benjamini and Hochberg 1995) controlled the false discovery rate (FDR) at 0.05 as the primary adjustment, while a Benjamini-Yekutieli adjustment (Benjamini and Yekutieli 2001) served as a sensitivity analysis for correlated retrieval outcomes. We considered model differences statistically supported when the adjusted *p*-value fell below 0.05 and the absolute rank-biserial correlation reached at least 0.10 to prevent interpreting negligible differences as meaningful. Finally, we evaluated robustness with a participant-clustered bootstrap that resampled children while retaining all of their contributed recording-target queries to preserve observation dependence.

#### Longitudinal analyses

Our primary longitudinal estimand determined whether a participant’s average target-present Hit@10 differed across the nominal 14, 24 and 36-month assessments. For each participant, wave and model, we averaged the query-level outcomes across eligible target-present queries prior to model fitting. We designated Hit@10 for Jina Embeddings v5 Omni as the primary model-endpoint combination, treating analyses of other models, metrics, modalities and individual targets as secondary or exploratory.

To estimate population-average developmental variation, we utilized generalized estimating equations (GEEs) (Liang and Zeger 1986). We parameterized these models with a Gaussian family, identity link, exchangeable working correlation, robust sandwich standard errors and participant-level clustering. This primary analysis included the 94 participants evaluated at two or more waves. We modeled the developmental wave continuously based upon nominal age in months to yield an estimated mean change per month.

Subsequently, we evaluated differences in developmental trajectories among models by incorporating model, time and model-by-time interaction terms in a joint GEE. We assessed the interaction coefficients jointly utilizing a Wald test rather than comparing separately fitted model-specific coefficients. To determine whether the observed developmental patterns merely reflected shifts in target composition, we repeated these analyses twice. We first excluded child and maternal speech targets, and separately, we excluded eight infant paralinguistic targets. Finally, our sensitivity analyses included the 62 complete-case participants represented at all three waves, compositional ablations that excluded speech or infant-paralinguistic targets, a secondary all-query GEE and a joint model-by-time interaction test (Supplementary Table S6).

#### Validation of retrieval-derived developmental language measures

We evaluated three language measures computed from retrieved child-speech segments, specifically CVD, MLU and SWP. Consistent with Section 2.7, our primary validation utilized text-to-audio child-speech retrieval for Jina Embeddings v5 Omni and did not pool text-to-video rankings. For each metric, we compared the retrieval-derived values against complete-session reference values independently at 14, 24 and 36 months. We excluded recordings with undefined retrieval-derived values, which resulted in a 14-month sample size of n = 105 for CVD and n = 75 for both MLU and SWP. We selected Spearman correlation as the primary metric to quantify how accurately the retrieved subsets preserved the rank ordering of recordings from lowest to highest language complexity. We utilized mean absolute error (MAE) to capture the typical magnitude of disagreement and evaluated signed error to detect systematic biases, where negative values indicated that the retrieval underestimated the complete-session reference. Furthermore, we estimated within-wave 95% confidence intervals for the Spearman correlations utilizing 500 recording-level bootstrap resamples. Finally, we reported MAE and signed error as secondary descriptive outcomes.

#### General statistical procedures

All hypothesis tests were two-sided. We defined statistical significance as P < 0.05 after applying the applicable multiplicity adjustments. We did not impute missing observations. We omitted prospective sample-size calculations because our analyses strictly utilized all eligible recordings within the fixed archival dataset. All analyses were conducted in Python 3.11.9 using the NumPy (Harris et al. 2020), SciPy (Virtanen et al. 2020), pandas (McKinney 2010) and statsmodels (Seabold and Perktold 2010) packages.

#### Ethics Statement

This retrospective computational study used archived caregiver-child recordings and CHAT transcripts from the National Evaluation of Early Head Start corpus, distributed through the Child Language Data Exchange System, a component of TalkBank. The present investigators did not recruit participants or collect new data. TalkBank states that “TalkBank materials have already received IRB review at the contributor’s home institution and no further IRB approval is needed for use of these data.” The applicable TalkBank policy is available at https://talkbank.org/0share/access.html. Data were accessed and analyzed in accordance with the applicable TalkBank access requirements and Ground Rules, available at https://talkbank.org/0share/rules.html. Accordingly, this study was conducted as a secondary analysis of archived research data under the ethical and data-sharing framework established by the original investigators and TalkBank.

#### Data Availability

The archived caregiver-child video recordings and CHAT transcripts analyzed in this study are available through the National Evaluation of Early Head Start, Harvard-Brattleboro, Vermont site, Corpus within CHILDES and TalkBank. The corpus can be accessed at https://talkbank.org/childes/access/Eng-NA/EHS.html and is permanently identified by https://doi.org/10.21415/ESE3-M119. Access to the transcripts and associated media is administered by TalkBank and is subject to its registration requirements, Ground Rules, and Code of Ethics. The Ground Rules are available at https://talkbank.org/0share/rules.html. The source recordings and transcripts remain hosted by TalkBank and will not be redistributed by the authors. The analysis code and non-media outputs required to reproduce the reported findings will be deposited on Github upon publication.

#### Author contributions

B.M. conceived and designed the study, acquired the TalkBank data, performed data preprocessing, developed the analytical code, conducted the statistical analyses, and drafted and revised the manuscript. M.-J.W. contributed to data management, software development and code review, and manuscript preparation and revision. A.P., R.M., and G.A. contributed to study conceptualization and design, interpretation of the findings, and critical drafting and revision of the manuscript. All authors reviewed and approved the final manuscript.

## Results

### 3.1. Analytic sample and retrieval evaluation

The initial archive contained 279 wave-specific mother-child CHAT files. We excluded two 14-month files lacking processed recording directories, yielding a final cross-model analytical sample of 277 recordings from 121 participants (105 at 14 months, 94 at 24 months, and 78 at 36 months; **Fig. 1**). Temporal segmentation produced 20,195 consecutive, non-overlapping chunks up to 20 seconds long, totaling 111.45 hours of audiovisual data. We evaluated each recording against 27 prespecified targets, producing 7,479 recording-target queries per model and 22,437 total evaluations across LanguageBind, Wave7B, and Jina Embeddings v5 Omni. Ninety-four participants contributed recordings at two or more developmental waves, with 62 represented at all three waves to form the complete-case sensitivity panel.

### 3.2. Cross-model retrieval performance

Across 7,479 aligned recording-target queries, we evaluated phrase retrieval separately against stored audio embeddings (text-to-audio) and stored video embeddings (text-to-video). For text-to-audio, Jina Embeddings v5 Omni retrieved at least one target-positive chunk among the top ten candidates for 38.3% of queries, slightly outperforming LanguageBind (37.0%) and Wave7B (36.1%) (Fig. 2a). Jina simultaneously achieved the highest Precision@10 (0.159), Recall@10 (0.113), MRR (0.238), and nDCG@10 (0.185) (Fig. 2b-2e and Table 3). For text-to-video, Jina again led with a Hit@10 of 36.4%, outperforming Wave7B (35.0%) and LanguageBind (34.5%), and likewise recorded the highest Precision@10 (0.160), Recall@10 (0.108), MRR (0.227), and nDCG@10 (0.181).

**Fig. 2.**
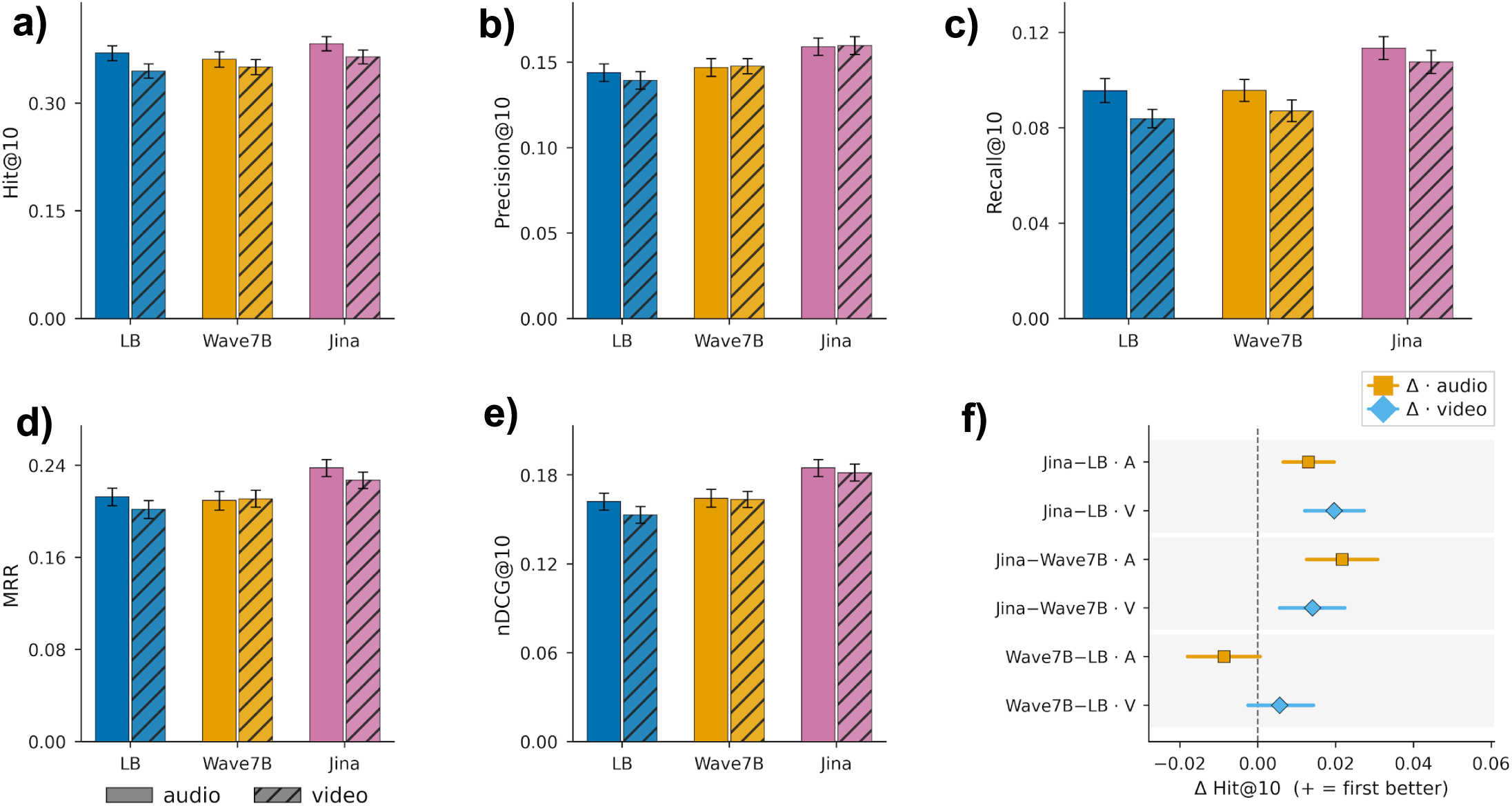
Combined phrase-based cross-modal retrieval performance. a-e, Retrieval performance across 7,479 aligned recording-target queries per model and retrieval channel for LanguageBind, Wave7B, and Jina Embeddings v5 Omni. For each model, solid bars show text-to-audio retrieval and hatched bars show text-to-video retrieval. Bars are query-weighted means for Hit@10, Precision@10, Recall@10, mean reciprocal rank (MRR) and normalized discounted cumulative gain at 10 (nDCG@10). Error bars indicate participant-clustered bootstrap 95% confidence intervals, with all repeated recordings and queries from each sampled participant retained. Hit@10 indicates whether at least one target-positive chunk occurred among the ten highest-ranked candidates. Precision@10 and Recall@10 quantify the proportion of retrieved and available target-positive chunks, respectively. MRR reflects the position of the first target-positive chunk, whereas nDCG@10 incorporates the ranked positions of all target-positive chunks. Higher values indicate better retrieval performance. f, Mean participant-level paired differences in Hit@10 with participant-clustered bootstrap 95% confidence intervals, shown separately for audio (A; orange squares) and video (V; blue diamonds) retrieve channels. The dashed vertical line denotes no difference, and positive values favor the first model named in each contrast. The 7,479 queries represent 277 recordings evaluated against 27 targets; each query was evaluated independently by all three models on both channels. LB, LanguageBind; MRR, mean reciprocal rank; nDCG, normalized discounted cumulative gain.

**Table 3.** Participant-level paired Hit@10 differences in the combined sample (dual retrieve channels)

| Retrieve channel | Comparison | Mean paired difference, $\Delta$ | 95% CI for $\Delta$ | Rank-biserial r | P<sub>BH</sub> |
| --- | --- | --- | --- | --- | --- |
| text-to-audio | Jina v5 Omni vs LanguageBind | 0.0140 | [0.0068, 0.0213] | 0.423 | < 0.001 |
| text-to-audio | Jina v5 Omni vs Wave7B | 0.0218 | [0.0118, 0.0323] | 0.443 | < 0.001 |
| text-to-audio | Wave7B vs LanguageBind | -0.0078 | [-0.0183, 0.0022] | -0.098 | 0.407 |
| text-to-video | Jina v5 Omni vs LanguageBind | 0.0172 | [0.0079, 0.0266] | 0.442 | < 0.001 |
| text-to-video | Jina v5 Omni vs Wave7B | 0.0117 | [0.0018, 0.0211] | 0.303 | 0.008 |
| text-to-video | Wave7B vs LanguageBind | 0.0055 | [-0.0049, 0.0166] | 0.112 | 0.348 |

**Note.** Paired Wilcoxon signed-rank tests on participant-level Hit@10 (n = 121 participants). Rows are model contrasts within each retrieve channel (text-to-audio; text-to-video). Positive $\Delta$ and rank-biserial r favour the first-named model. $P_{BH}$ is the Benjamini-Hochberg adjustment across the 15 aggregate tests within each channel (Hit@10 is one of those five metrics) . Full five-metric pairwise results, including $P_{BY}$ , are in Supplementary Table S5.

Participant-level paired analyses supported Jina’s Hit@10 advantage in both retrieve channels (n = 121; Table 3). For text-to-audio retrieval, mean paired difference Hit@10 was 0.0140 versus LanguageBind (rank-biserial r = 0.423; *P*_BH < 0.001) and 0.0218 versus Wave7B (r = 0.443; *P*_BH < 0.001). On the other hand, for text-to-video retrieval, the corresponding advantages were 0.0172 versus LanguageBind (r = 0.442; *P*_BH < 0.001) and 0.0117 versus Wave7B (r = 0.303; *P*_BH = 0.008). All four contrasts remained significant after Benjamini-Yekutieli adjustment (Supplementary Table S5). Participant-clustered bootstrap 95% confidence intervals excluded zero (Fig. 2f).

Descriptively, Jina had the highest all-query Hit@10 at every developmental wave in both retrieve channels (Fig. 3a). For text-to-audio retrieval, Jina Hit@10 was 0.399, 0.394 and 0.347 at 14, 24 and 36 months, respectively; margins over the second-best model were 0.004, 0.015 and 0.012. For text-to-video retrieval, the corresponding values were 0.362, 0.388 and 0.339, with margins of 0.003, 0.027 and 0.009. Paired, participant-clustered contrasts support a Jina advantage in most wave×channel comparisons, although a minority of Jina−Wave7B (video) intervals are compatible with no difference (Fig. 3b). Cross-wave differences in absolute Hit@10 should not be read as uniform encoder degradation with child age; Figure 4 shows that those level shifts are largely compositional. We evaluate developmental dynamics with repeated-measures longitudinal models in Section 3.4. Between the comparator models, absolute ranking was channel- and metric-dependent; the participant-level Wave7B−LanguageBind difference in Recall@10 was not significant after multiplicity adjustment (Supplementary Table S5). Collectively, these absolute retrieval rates position phrase-based search as an effective tool for prioritizing candidate evidence for expert review, rather than a standalone replacement for behavioral coding.

**Fig. 3.**
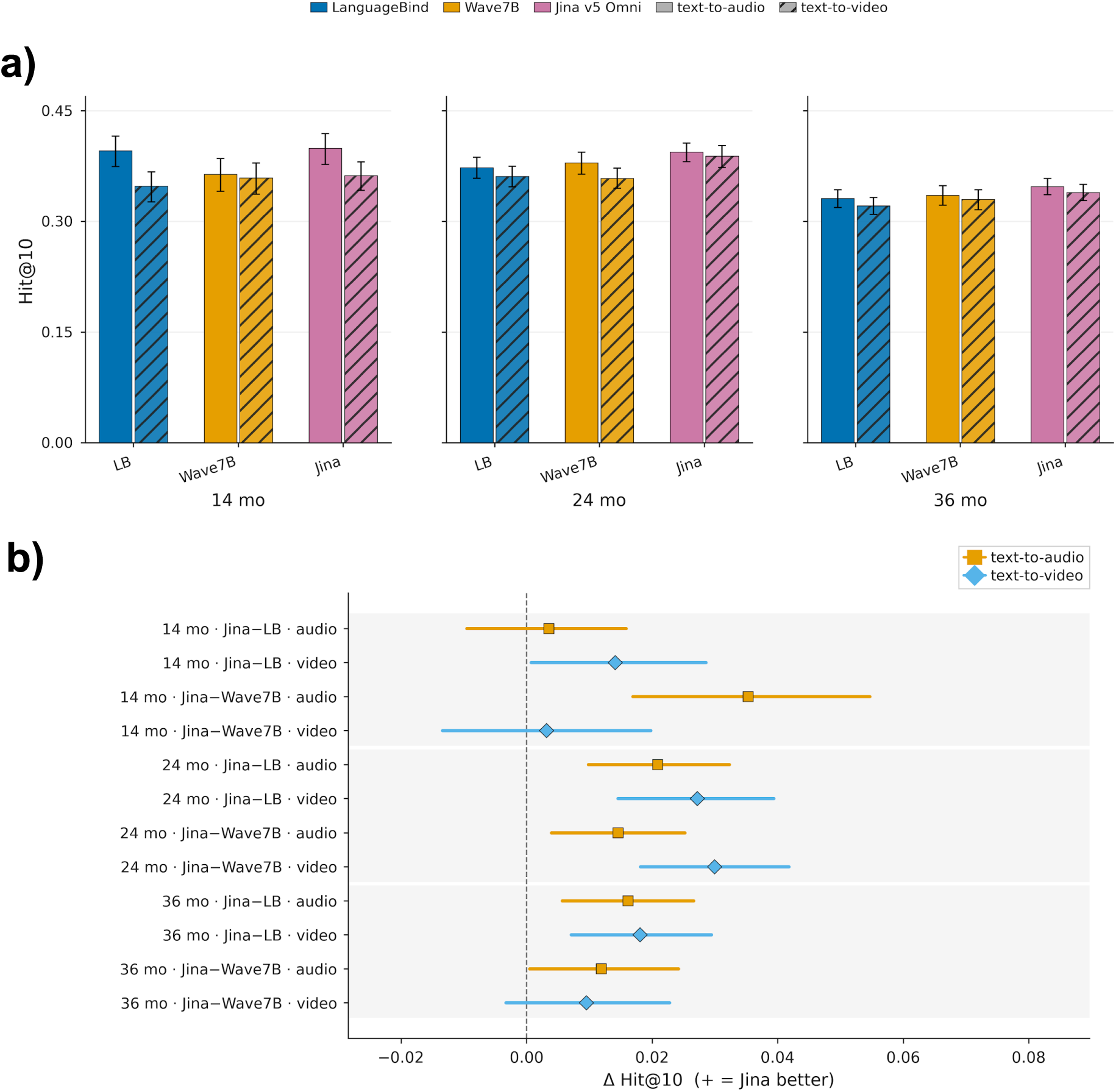
Wave-stratified Hit@10 levels and stable model ranking across retrieve channels. Top: all-query Hit@10 (± bootstrap 95% CI) at 14, 24 and 36 months, each wave drawn as its own panel (no connecting age trajectory). Colour encodes model; solid bars = text-to-audio, hatched bars = text-to-video. Bottom: participant-clustered bootstrap 95% confidence intervals for paired Δ Hit@10 (Jina v5 Omni − LanguageBind; Jina − Wave7B) within each wave and channel. Orange squares = text-to-audio; blue diamonds = text-to-video; values to the right of zero favour Jina. Jina remains the leading or tied encoder at every wave: most contrasts exclude zero, while a minority of Jina−Wave7B video comparisons are compatible with no difference (confidence interval crosses zero). Differences in absolute Hit@10 across the top panels reflect changing available behaviours in the corpus, not a uniform loss of encoder quality with child age that compositional mechanism is shown in Figure 4. Sample sizes: text-to-audio: 14 months n=2835, 24 months n=2538, 36 months n=2106; text-to-video: 14 months n=2835, 24 months n=2538, 36 months n=2106

**Fig. 4.**
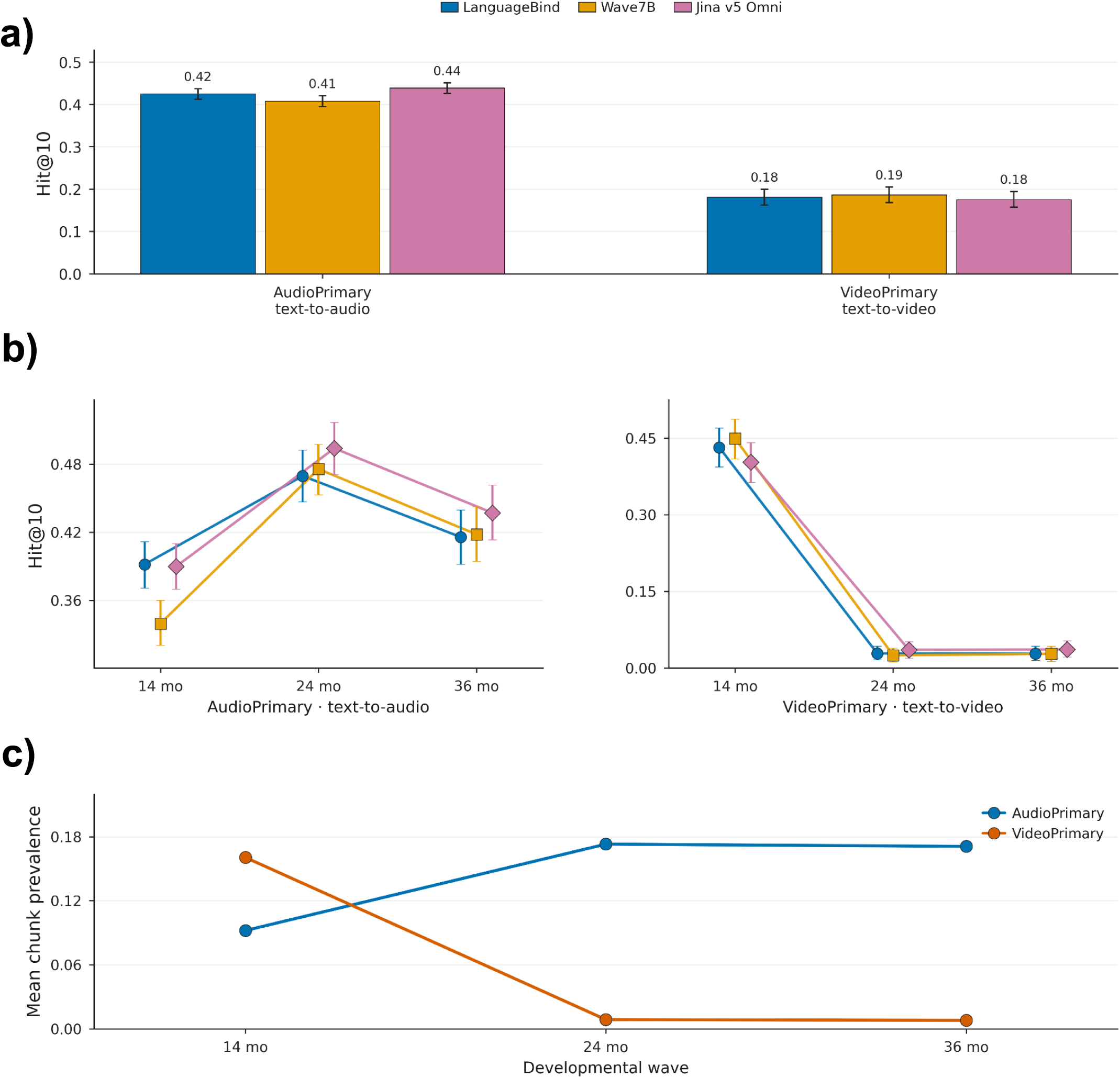
Domain-matched retrieval and the compositional collapse of VideoPrimary Hit@10 after 14 months. a) combined-cohort Hit@10 (± bootstrap 95% CI) for domain-matched retrieve only: AudioPrimary targets via text-to-audio; VideoPrimary targets via text-to-video. Within AudioPrimary text-to-audio, Jina v5 Omni is highest (LanguageBind 0.42, Wave7B 0.41, Jina v5 Omni 0.44). Within VideoPrimary text-to-video, models are nearly tied (LanguageBind 0.18, Wave7B 0.19, Jina v5 Omni 0.18; bootstrap 95% CIs overlap). b) The same matched estimands by developmental wave. AudioPrimary text-to-audio Hit@10 does not fall monotonically (Jina: 0.39 to 0.49 to 0.44). VideoPrimary text-to-video falls sharply (Jina: 0.40 to 0.04 to 0.04), but this is not age-related encoder degradation. c), Mean chunk prevalence by taxonomy shows the mechanism: VideoPrimary content nearly vanishes after 14 months, whereas AudioPrimary prevalence rises then plateaus. Three structured activities three bags, frustration task and teaching task, account for 98% of the VideoPrimary Hit@10 drop from 14 to 24 months (e.g. three bags: prevalence 0.51 to 0.02, Jina Hit@10 0.95 to 0.02); frustration and teaching reach zero prevalence (and zero Hit@10) at 24-36 months. Because all-query Hit@10 requires a target-positive recording, absent behaviours force Hit@10 toward zero even if the encoder is unchanged. Pointing is the VideoPrimary flag that remains present and does not decline from 14 to 24 months (Jina Hit@10 0.13 to 0.18). Any apparent all-query age decline is therefore largely compositional (changing available behaviours), not a uniform encoder failure with child age.

### 3.3. Retrieval varies by target domain and developmental composition, not by uniform encoder decline with age

Domain-matched Hit@10 differed sharply by target taxonomy (Fig. 4a). For AudioPrimary targets retrieved through text-to-audio, combined-cohort Hit@10 was 0.44 (Jina), 0.42 (LanguageBind) and 0.41 (Wave7B). For VideoPrimary targets retrieved via text-to-video, the same models were much lower and nearly tied (0.18, 0.18 and 0.19). The same matched estimands by developmental wave show that this is not age-related model failure (Fig. 4b): AudioPrimary text-to-audio Hit@10 did not fall monotonically (Jina: 0.39, 0.49, 0.44 at 14, 24 and 36 months), whereas VideoPrimary text-to-video collapsed after 14 months (Jina: 0.40, 0.04, 0.04). Mean chunk prevalence by taxonomy explains the collapse (Fig. 4c), VideoPrimary content nearly vanished after 14 months, while AudioPrimary prevalence rose then plateaued. At the flag level three structured activities three bags, frustration task and teaching task, account for 98% of the VideoPrimary Hit@10 drop from 14 to 24 months (e.g. three bags: prevalence 0.51 to 0.02, Jina Hit@10 0.95 to 0.02); frustration and teaching reach zero prevalence at 24-36 months. Because all-query Hit@10 requires a target-positive recording, absent behaviours force Hit toward zero even if ranking quality is unchanged. Therefore, “pointing” the VideoPrimary flag that remains present did not decline from 14 to 24 months (Jina Hit@10 0.13 to 0.18). For text-to-audio retrieval with Jina, Hit@10 was highest for maternal speech (0.986), maternal questions (0.971), observer speech (0.903), child speech (0.888) and laughing (0.812), and among the lowest for babbling (0.087), whimpering (0.047), shushing (0.018) and smiling (0.007). LanguageBind and Wave7B showed the same easy-versus-hard ordering, indicating that difficulty is largely target-driven (Supplementary Figs. S1 and S2). Chunk prevalence spanned nearly three orders of magnitude (0.785 for maternal speech to 0.000938 for shushing) and tracked Hit@10 (Jina Spearman ρ = 0.93).

Because a recording may simply lack a given behaviour, we also restricted analysis to the 3,801 recording-target queries with at least one annotated positive chunk and split those present-target queries by chunk prevalence into common (≥5%) and rare (<5%) strata. In text-to-audio, the 2,834 common-target queries were often recovered by every model (68% all-model hits; 7% all-model misses), whereas the 967 rare-target queries were much harder (28% all-model hits; 28% all-model misses); text-to-video showed the same pattern (common: 64% / 8%; rare: 21% / 33%). Rare targets were therefore harder even when annotated evidence was present, so target absence alone cannot explain the full performance drop.

### 3.4 Target-present Hit@10 improved with age; apparent all-query declines were prevalence-confounded and composition-dependent

Across the full 27-target panel, all-query Jina Hit@10 fell from 0.399 at 14 months to 0.394 at 24 months and 0.347 at 36 months under text-to-audio retrieval (text-to-video: 0.362, 0.388, and 0.339; Fig. 3). Because target-absent queries score zero by definition and the mix of available behaviors shifts across waves, these descriptive means conflate retrieval skill with developmental change in target presence. Because absent targets yield Hit@10 = 0 by definition, we therefore ran the primary longitudinal GEE only on target-present queries, asking whether the model can find the behavior when it is actually in the recording, using the 94 participants seen at two or more waves and pooling text-to-audio with text-to-video. Under that definition, Jina Hit@10 rose with age: β = 0.00299 per nominal month (robust 95% CI, 0.00151 to 0.00446; *P* < 0.001), or about +0.066 from 14 to 36 months (Fig. 5a). The same positive pattern held in the 62 complete-case participants (β = 0.00303 per nominal month; 95% CI, 0.00132 to 0.00474; *P* < 0.001). A secondary GEE that put target-absent queries back in still on those 94 participants flipped the sign (β = -0.00197 per nominal month; 95% CI, -0.00294 to -0.00100; *P* < 0.001). In short, all-query age slopes look like decline mainly because target presence changes with age and therefore when the target is present, retrieval improves.

**Fig. 5.**
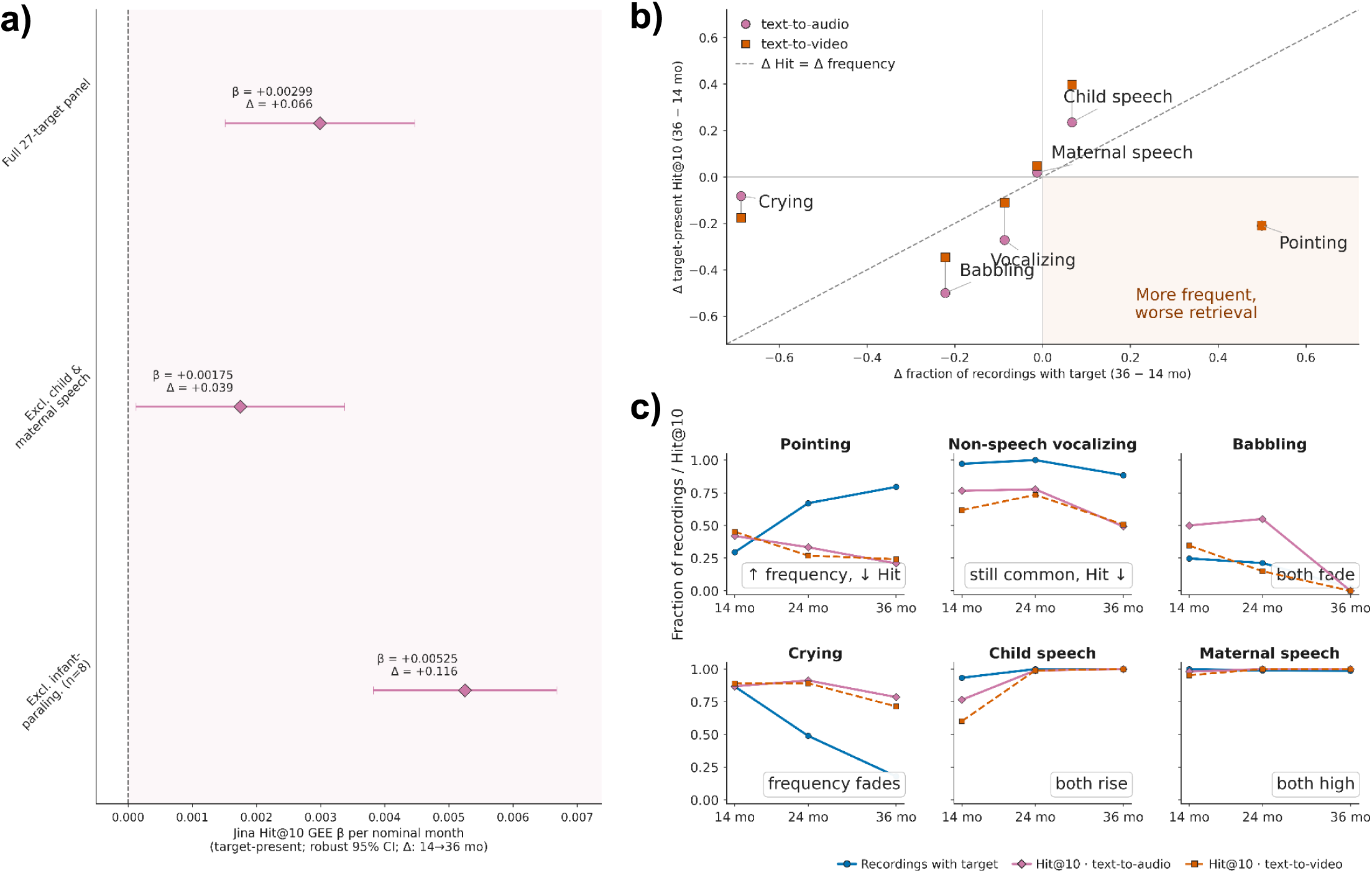
Target-present age gains depend on target composition, and target frequency does not track Hit@10. **a)** Jina v5 Omni Hit@10 GEE slope per nominal month (robust 95% CI) on target-present queries among participants assessed at two or more waves (n = 94; text-to-audio and text-to-video retrieve channels pooled). Printed values give the point estimate β and the mapped absolute change Δ = β × 22 months between the 14- and 36-month waves. Relative to the full 27-target panel, excluding child and maternal speech attenuates the positive slope, whereas excluding eight infant-paralinguistic targets (babbling, crying, gasping, laughing, screaming, non-speech vocalizing, whimpering, and whining) steepens it. b) Change from 14 to 36 months in the fraction of recordings containing each target (x) versus target-present Hit@10 (y) for six exemplar behaviors. Circles, text-to-audio; squares, text-to-video. The dashed diagonal is the equal-change (mirror) reference; points off this line indicate that frequency shifts do not track retrieval shifts. The shaded lower-right region marks behaviors that became more frequent while Hit@10 fell (notably pointing). c) Wave-wise trajectories for the same targets on a shared 0-1 axis: blue, fraction of recordings in which the target is present; pink/orange, Jina target-present Hit@10 under text-to-audio / text-to-video. Nominal wave ages of 14, 24, and 36 months were used in continuous-wave models and should not be read as each participant’s exact chronological age.

Compositional ablations changed the magnitude but not the direction of the target-present age gain (Fig. 5a). Excluding child and maternal speech attenuated the slope to 0.00175 per nominal month (95% CI, 0.00012 to 0.00338; *P* = 0.035; mapped Δ = 0.039). Excluding eight infant-paralinguistic targets (babbling, crying, gasping, laughing, screaming, non-speech vocalizing, whimpering, and whining) steepened it to 0.00525 per nominal month (95% CI, 0.00382 to 0.00668; *P* < 0.001; mapped Δ = 0.116). Thus early affective/vocal targets are not the sole drivers of a systemic collapse; removing them reveals a larger age-related gain on the remaining panel (Supplementary Table S6).

Longitudinal target frequencies also did not mirror retrieval success (Fig. 5 b and c). Babbling and crying declined in both prevalence and Hit@10. Non-speech vocalizing remained common at 36 months yet became substantially harder to retrieve. Pointing became more frequent while target-present Hit@10 fell. Finally, a model-by-time interaction was present but modest (Wald χ²(4) = 10.00, *P* = 0.040): target-present Hit@10 improved with age for Jina, LanguageBind, and Wave7B, with model-specific rates of gain rather than a universal age-related retrieval limit.

### 3.5 Text-to-audio retrieved segments preserved relative language differences but not complete-session values

Because child vocabulary diversity (CVD), mean length of utterance (MLU), and single-word proportion (SWP) are properties of the speech stream, we restricted developmental language validation to text-to-audio retrieval of child-speech queries and did not pool text-to-video rankings. Within each developmental wave, language metrics computed from Jina Embeddings v5 Omni top-ranked child-speech segments correlated positively with complete-session human-coded baselines (Fig. 6a). For CVD, Spearman ρ was 0.68, 0.82, and 0.90 at 14, 24, and 36 months, respectively. Corresponding correlations were 0.74, 0.84, and 0.84 for MLU, and 0.77, 0.81, and 0.78 for SWP. CVD rank agreement was hardest at 14 months where many sessions have sparse or tied vocabularies and strengthened at later waves, arguing against a simple age-related collapse in relative recovery. Preserving within-wave rank order did not, however, yield accurate absolute estimation (Fig. 6b). CVD showed severe, systematic underestimation: Jina’s mean absolute error rose from 3.5 unique words at 14 months to 64.9 at 24 months and 96.7 at 36 months, and mean signed errors were negative at every wave (no recording was overestimated). Retrieved segments therefore identified children with relatively larger vocabularies within age while capturing a rapidly diminishing fraction of total lexical output as sessions grew richer. MLU and SWP mean signed errors remained small, with modest MAEs (MLU: 0.17-0.36 words/utterance; SWP: 0.08-0.10), indicating that utterance-structure summaries are more stably recovered from top-ranked windows than absolute vocabulary tallies. At 14 months, MLU and SWP have n=75 (vs CVD n=105) because recordings without eligible child utterances in the text-to-audio retrieved subset were excluded. We conclude that text-to-audio search can locate speech-dense windows that stratify developmental language variation within age, but cannot currently substitute for complete-session linguistic measurement.

**Fig. 6.**
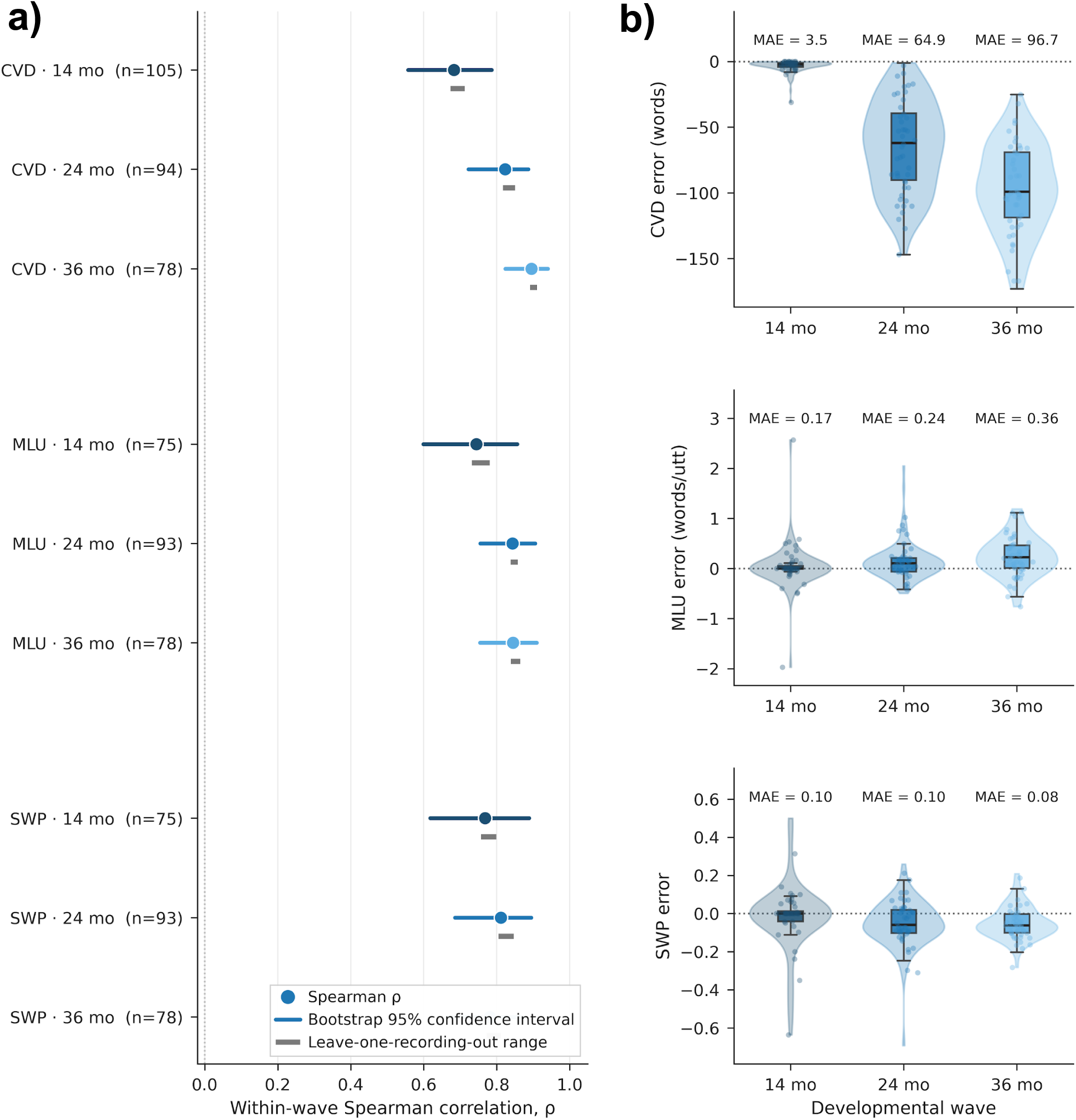
Text-to-audio child-speech retrieval preserves within-wave language ranks, but absolute vocabulary size is under-recovered as sessions grow richer. All estimates use Jina Embeddings v5 Omni text-to-audio top-ranked “child speech” segments only language measures are not derived from text-to-video rankings. **a)** Within-wave Spearman ρ between retrieval-derived and complete-session human-coded child vocabulary diversity (CVD), mean length of utterance (MLU), and single-word proportion (SWP). Points = ρ; coloured lines = recording-level bootstrap 95% CIs; gray bars = leave-one-recording-out range. **b)** Signed errors (retrieval - complete-session reference) for the same measures, with MAE annotated above each wave. The left panel asks whether retrieval keeps children in the right order within age, while the right panel asks whether the retrieved subset recovers absolute session values.

## Discussion

In this retrospective evaluation of 277 Early Head Start recordings, prespecified natural-language phrases successfully retrieved temporally localized developmental behaviors directly from audio and video streams, entirely bypassing transcript text. Although Jina Embeddings v5 Omni achieved the highest overall performance, its text-to-audio top-10 success rate of 38.3% (text-to-video 36.4%) indicates that zero-shot retrieval remains incomplete. Performance depended heavily on the queried behavior. The models reliably retrieved common speech targets but consistently failed on rare vocal, affective and gestural events. Furthermore, apparent all-query Hit@10 declines with age were largely compositional, driven by disappearing VideoPrimary activities and shifting target presence, whereas target-present Hit@10 improved with age for all three models. While retrieval-derived child speech segments preserved within-wave rank order for language metrics, they systematically under-recovered absolute vocabulary size. Consequently, we position phrase-based retrieval as an evidence-selection layer to prioritize candidate moments for human review rather than a standalone replacement for behavioral coding or language assessment.

These findings shift the computational objective from assigning predefined behavioral labels to localizing the raw audiovisual evidence required for expert judgment. Prior systems have shown that developmental audiovisual recordings can support the automated measurement of affect and attention, caregiver responsiveness, engagement states, stereotyped motor behavior and infant movement patterns (Barami et al. 2024; Groos et al. 2022; Egger et al. 2018; Isaev et al. 2024; Withanage Don et al. 2024) . However, these systems generally estimate a prespecified construct selected during model development or reduce the recording to a derived score, class or event count. Our results establish a complementary paradigm. A single indexed recording can be queried for multiple prespecified auditory and visual concepts, returning segments that retain their original interactional context. This distinction is critical because isolated event detection inherently strips away developmental meaning. A vocalization or gesture becomes clinically informative only when analyzed alongside its timing, antecedents and dyadic consequences. Phrase-based retrieval therefore offers a highly scalable intermediate layer that narrows lengthy recordings to inspectable moments before manual measurement. Nevertheless, the moderate retrieval rates and severe target-specific variations confirm that this flexibility does not equate to unrestricted behavioral understanding.

Among the tested models, Jina led all five retrieval metrics and returned relevant segments earlier and more densely within the top ten results (Fig. 2a-2e and Table 3). This consistent superiority across paired comparisons and retrieval depths indicates a robust algorithmic advantage. Absolute performance, however, remained modest. For Jina, Precision@10 and Recall@10 were 0.159 and 0.113 for text-to-audio, and 0.160 and 0.108 for text-to-video. This indicates that most returned candidates lacked the target behavior and the model recovered only a limited fraction of available positive segments. Moreover, on the primary target-present estimand, Hit@10 improved with age for Jina, LanguageBind, and Wave7B, demonstrating a modest model-by-time interaction (Wald χ²(4) = 10.00, *P* = 0.040). For Jina, excluding child and maternal speech attenuated this gain, whereas excluding eight infant-paralinguistic targets steepened it. This demonstrates that early affective and vocal targets are not the sole drivers of a collapse, and removing them reveals a larger age-related gain on the remaining panel. Apparent all-query age declines reverse sign once target-absent queries are restored, which confirms prevalence confounding rather than uniform encoder failure. We conclude that while embedding models differ materially in ranking capabilities, their success remains strictly contingent on the behavioral target and the developmental context. Related systems have used brief elicitation protocols, home-recorded task videos, caregiver screening instruments, and voice features to classify autism risk or symptom severity (Kim et al. 2025; Perochon et al. 2023; Bae et al. 2025). These studies address a clinically important but distinct task involving classification or risk stratification under dedicated protocols rather than the localization of prespecified behaviors within archival recordings.

Model differences operated beneath a dominant target-level performance gradient. Across all architectures, target prevalence strongly correlated with retrieval success (Jina Spearman ρ = 0.93 for text-to-audio, Fig. 4). Operationally, models returned common speech targets far more frequently than rare vocal, affective, and gestural targets, including pointing, babbling, whimpering, and smiling. The ubiquity of this failure pattern suggests that simply swapping general-purpose embedding models will not resolve the underlying limitation. Several factors likely contribute to this bottleneck. Rare targets present fewer positive training windows, and brief events easily dilute within fixed 20-second segments. Furthermore, infant vocalizations frequently suffer from acoustic masking by caregiver speech or background noise, while gestures frequently occur off-camera or under occlusion. In addition, general-purpose pretraining datasets likely used in present models vastly underrepresent these fleeting, developmentally specific behaviors, creating a severe domain shift (Radford et al. 2021; Birhane et al. 2021; Schmarje 2024). Future evaluations must therefore incorporate speaker-matched hard negatives, semantically related distractors, and expert review of concept specificity to untangle these error sources. Because pointing and socially contingent vocalizations are foundational to early communication (Goldstein et al. 2003; Rowe and Goldin-Meadow 2009; Warlaumont et al. 2014), the current system proves unreliable for the most developmentally critical behaviors.

This target dependence heavily influences the observed developmental patterns. In our primary longitudinal analysis, target-present Hit@10 for the Jina model rose across the nominal 14, 24 and 36-month assessments. Removing eight infant-paralinguistic targets did not erase this association but rather strengthened it, whereas speech ablation attenuated it. Apparent all-query declines are therefore driven by prevalence and composition rather than a uniform deterioration in audiovisual processing. This distinction matters deeply because early communication changes rapidly over this window. A single query phrase must span acoustically and visually divergent manifestations as prelinguistic vocalizations evolve into complex speech (Brown 2009; Fenson and Others 2007). Consequently, we interpret these longitudinal findings as evidence of developmental nonstationarity rather than a simple age effect. Future research must examine target-specific trajectories, verify that query semantics remain invariant across age and evaluate models trained on developmentally diverse samples. Developmental retrieval systems require validation not merely for static accuracy but for their capacity to adapt as a child’s interactional repertoire matures. In autistic preschoolers, automated acoustic analyses similarly showed that associations between prosodic features and developmental phenotypes differed by language-production stage, although some voice-quality features remained comparatively stable across expressive levels (Godel et al. 2023). These constraints strictly bounded the language analyses. Retrieval-derived vocabulary diversity, mean length of utterance and single-word proportion preserved substantial recording-level ordering, with Jina demonstrating the strongest rank correlations. However, rank correlation does not imply accurate complete-session recovery. Vocabulary diversity functions cumulatively, and thus ten retrieved segments necessarily captured a diminishing fraction of the child’s expanding lexical repertoire. Several language-sample measures are sensitive to transcript length, which reinforces the need to interpret retrieval-derived values as subset estimates rather than complete-session measurements (Heilmann et al. 2010). Consequently, while Jina preserved high within-wave correlations for vocabulary diversity using text-to-audio child-speech retrieval, absolute recovery ultimately failed. Specifically, the mean absolute error increased from 3.5 unique words at 14 months to 64.9 words at 24 months and reached 96.7 words at 36 months. Therefore, although retrieved segments can effectively stratify children within a specific age group, they cannot substitute for a complete-session lexical measurement.

Given these boundaries, the most credible near-term clinical application of these multimodal models is retrieval-assisted review. Phrase-based search can render massive audiovisual archives tractable by directing coders to candidate moments of speech, affect or gesture, preserving the source segment for expert interpretation. This workflow supports secondary analyses, consensus review and the curation of rare-event examples for active learning. Medium-term applications such as fidelity review in caregiver-mediated interventions or indexed longitudinal tracking for remote specialists remain plausible but demand task-specific prospective validation. Crucially, the translational value of this framework cannot rest on retrieval accuracy alone. Future reader studies must determine whether ranked clips genuinely reduce expert review time without inflating missed events, false-positive burdens or interpretive errors (Vasey et al. 2022). Evaluation must prioritize interrater reliability, cognitive workload and the risk of automation bias when models present plausible but incorrect evidence (Lyell and Coiera 2017). Finally, developmental AI systems must be engineered to quantify their own statistical uncertainty and automatically withhold predictions when evidence is insufficient (Megerian et al. 2022). For rare targets or low-confidence searches, outputting an explicit insufficient-evidence warning is clinically safer than presenting a ranked list that invites unwarranted interpretation. The present results establish technical feasibility for evidentiary prioritization while strictly prohibiting autonomous screening, diagnosis or complete-session assessment. Several design features strengthen the internal validity of this benchmark, including a large longitudinal sample of 111.45 hours from 277 recordings, identical cross-model candidate pools and the strict exclusion of transcript text from the retrieval channel. Furthermore, the source corpus is actively maintained (MacWhinney 2007; Pan, B. A., Ayoub C., Snow, C. E. 2008) and we utilized publicly documented open-weight multimodal embedding models(Zhu et al. 2023; Tang et al. 2025; Hönicke et al. 2026) to maximize reproducibility.

## Limitations

These strengths must be weighed against important limitations in the archival data and the evaluated developmental constructs. The corpus originated from a single site serving predominantly English-speaking, low-income families in rural New England and featured a largely European American source cohort (Pan et al. 2005). The absence of individual-level demographic data for our exact analytical sample precludes a formal evaluation of subgroup fairness or demographic transportability. This limitation is especially consequential in pediatric AI, as geographically, socioeconomically, and clinically unrepresentative datasets can undermine model validity and equitable deployment (Muralidharan et al. 2024). Because the recordings predominantly capture mother-child interactions during semi-structured activities, their generalization to unrestricted daylong behavior or interactions with other caregivers remains uncertain. Naturalistic sensing research further shows that infant behavior varies substantially across hours, routines, interaction contexts, and individuals. Consequently, no brief observation can be assumed to represent the true distribution of everyday experience (de Barbaro and Fausey 2022; Kaur et al. 2026). Furthermore, the source media date from the late 1990s and early 2000s (Pan, B. A., Ayoub C., Snow, C. E. 2008) and therefore, performance on contemporary video could diverge significantly owing to differences in compression, frame rate, camera placement, and background activity.

Construct validity presents a related limitation. The 27 targets reflect behaviors available in archived CHAT annotations rather than a prospectively designed developmental battery. Recent video-based studies centered on joint-attention tasks and child-play interactions illustrate how such constructs can be operationalized. However, these studies also demonstrate that measurement depends heavily on protocol-specific elicitation, annotation, and clinical reference standards (Choudhury et al. 2026). Furthermore, holding the natural-language query constant was necessary for controlled algorithmic comparisons, yet this approach does not confirm robustness to alternative phrasing. Because semantically equivalent prompts can generate disparate rankings in multimodal architectures (Mwangi et al. 2026; Jabbar Abd Sattar Hamoudi et al. 2026; Wang et al. 2024; Stewart et al. 2024), future research must systematically evaluate synonymous descriptors, age-conditioned formulations, and clinician-generated wording.

Technically, the fixed 20-second temporal windows potentially diluted brief vocalizations, combined disparate behaviors, and obscured precise event boundaries. The evaluation also restricted searches to known recordings, thereby testing within-session temporal localization rather than cross-corpus retrieval. Furthermore, standard GEE missingness assumptions (Robins et al. 1995) constrain our longitudinal statistical inferences. Finally, relying on three general-purpose, open-weight models without domain-specific fine-tuning effectively characterizes zero-shot transfer limits. It does not represent the theoretical upper bound of developmental retrieval performance.

## Future work and conclusion

Future work must prioritize domain adaptation, clinically targeted constructs, and prospective validation over the incremental optimization of this baseline benchmark. The immediate scientific priority is fine-tuning multimodal models utilizing developmentally representative recordings enriched for rare behaviors such as early gestures and affective signals. Second, the field must transition from isolated-event retrieval to dyadic sequence modeling. Because developmental meaning resides heavily in temporal contingency (Goldstein et al. 2003; Warlaumont et al. 2014; Cheung et al. 2021), modeling how caregiver responses shape child vocalizations will align computational targets directly with developmental theory. Finally, external validation must incorporate diverse clinical populations, varied linguistic contexts, and contemporary recording devices. This must occur alongside prespecified audits for algorithmic fairness, failure modes, and biometric data privacy.

In conclusion, prespecified phrase-based cross-modal retrieval successfully renders lengthy caregiver-child recordings searchable by localizing audiovisual evidence directly linked to the original interaction. While open-weight multimodal embeddings support inspectable evidence selection, their success remains heavily constrained by target prevalence, developmental expression, and the availability of dense behavioral signals. Although retrieved segments preserve recording-level language hierarchies, they currently offer no measurable advantage over child-speech-matched transcript subsets for independent language assessment. Consequently, phrase-based cross-modal search represents a technically feasible foundation for evidentiary prioritization. Its ultimate clinical utility will depend on improving the recovery of rare developmental behaviors, preserving expert human judgment, and embedding robust safeguards for privacy and algorithmic fairness.

## Acknowledgments

This work was supported by the NECMHR01 grant from the Texas Child Mental Health Care Consortium (TCMHCC) and The National Institute of Health (NIH) AIM-AHEAD consortium

