## Supplemental materials for "Natural-language retrieval with multimodal embeddings identifies candidate developmental behaviors in caregiver-child recordings"

**Supplementary Information**

**Data provenance, analytic sample, and observational structure.** Exclusion criteria strictly follow the parameters defined in the main text (Section 2.2). For longitudinal analyses, we linked recordings utilizing the corpus participant identifiers to account for repeated within-participant observations.

**CHAT parsing, temporal segmentation, and media extraction.** As outlined in the main text (Section 2.3), we partitioned recordings into 20-second chunks. To execute this, we utilized FFmpeg to extract audio as mono 16-bit pulse-code modulation at 16 kHz using the specific flags -vn -acodec pcm_s16le -ar 16000 -ac 1. We generated video-only streams utilizing the -an -c:v copy options. During transcript parsing, we strictly maintained utterance integrity; we did not divide utterances crossing a chunk boundary. We assigned untimestamped utterances to the most recently resolved chunk in transcript order, or to the initial chunk if no preceding timestamp existed. We retained chunks lacking verbal speech to ensure the preservation of observer speech, paralinguistic vocalizations, gestures, affective behavior, and visual activity contexts.

**Reference-label construction and excluded variables**

We deterministically derived chunk-level reference labels from the archived CHAT annotations, applying identical rules across all three embedding models. The final retrieval panel contained 27 targets (21 AudioPrimary and 6 VideoPrimary). We explicitly excluded TextPrimary variables and annotation-tier indicators (Supplementary Table S1) because their operational definitions represent properties of transcript text or annotation structures rather than audiovisual phenomena. Querying such variables utilizing natural-language text risks evaluating lexical correspondence rather than true cross-modal retrieval.

| **Category** | **Flag** | **Operational definition** |
| --- | --- | --- |
| TextPrimary | has_chi_zero_utterance | Child produced at least one zero or nonverbal utterance |
| TextPrimary | has_mot_zero_utterance | Mother produced at least one zero utterance |
| TextPrimary | has_chi_exclamation | Child utterance with an exclamation terminator |
| TextPrimary | has_mot_exclamation | Mother utterance with an exclamation terminator |
| TextPrimary | has_chi_incomplete | Child utterance with an incomplete terminator |
| TextPrimary | has_mot_incomplete | Mother utterance with an incomplete terminator |
| TextPrimary | has_chi_trailing | Child utterance with a trailing-off terminator |
| TextPrimary | has_mot_trailing | Mother utterance with a trailing-off terminator |
| TextPrimary | has_error_marker | Transcript contained error-coding markers |
| TextPrimary | has_retracing | Transcript contained retracing or reformulation markers |
| TextPrimary | has_uncertain | Transcript contained uncertain-transcription markers |
| TextPrimary | has_unintelligible | Transcript contained unintelligible material such as xxx |
| TextPrimary | has_chi_single_word_only | All verbal child utterances in the chunk contained one word |
| TextPrimary | has_mot_wh_question | Mother produced a wh-question |
| TextPrimary | has_mot_yn_question | Mother produced a yes-no question |
| TextPrimary | has_mot_imperative | Mother produced an imperative utterance |
| TextPrimary | has_mot_reading_aloud | Mother was coded as reading aloud |
| TextPrimary | has_postcode_pi | The CHAT pointing-included postcode was present |
| Annotation tier | has_mor | Morphology tier, %mor |
| Annotation tier | has_gra | Grammatical-relations tier, %gra |
| Annotation tier | has_act | Action tier, %act |
| Annotation tier | has_gpx | Gestural-proxemic tier, %gpx |
| Annotation tier | has_com | Comment tier, %com |
| Annotation tier | has_spa | Speech-act tier, %spa |
| Annotation tier | has_xpho | Phonology tier, %xpho |
| Annotation tier | has_tim | Timing tier, %tim |
| Annotation tier | has_par | Paralinguistic tier, %par |
| Annotation tier | has_sit | Situation tier, %sit |
| Annotation tier | has_exp | Explanation tier, %exp |

**Supplementary Table S1. CHAT-derived variables excluded from the phrase-retrieval target panel**. We excluded 18 TextPrimary variables and 11 annotation-tier indicators from phrase-retrieval targeting, retaining their labels exclusively for transcript auditing and secondary analyses.

**Lexical sanitization before transcript indexing**

Although we strictly excluded transcript text from the retrieval channel during phrase-only evaluation, we stored it as an auxiliary indexed representation. To prevent label leakage during secondary text analyses, we removed inflected forms of behavior-label stems prior to transcript indexing (Supplementary Table S2). This case-insensitive sanitization step included common morphological variants. Crucially, this process did not alter the audio, video, query phrases, or CHAT-derived reference labels.

| **Reference flag** | **Removed lexical forms** |
| --- | --- |
| has_crying | cry, cries, cried, crying, cries and squirms |
| has_laughing | laugh, laughs, laughed, laughing |
| has_pointing | point, points, pointed, pointing |
| has_babbling | babble, babbles, babbled, babbling |
| has_whimpering | whimper, whimpers, whimpered, whimpering |
| has_vocalizing | vocalize, vocalizes, vocalized, vocalizing, grunt, grunts, noise, noises, sound, sounds |
| has_overlap | overlap, overlaps, overlapping |
| has_pause | pause, pauses, pausing |

**Supplementary Table S2. Lexical sanitization blocklist.** This blocklist corresponds to the sanitization call executed by the indexing script. We did not include speaker-presence phrases (such as “Child speech”) in this blocklist because they do not share direct lexical stems with standard transcript content.

**Audiovisual indexing and retrieval infrastructure**

As detailed in the main text (Section 2.4), we ingested aligned audio, video, transcript text, and metadata through a unified gateway. Query execution strictly evaluated chunks from the same recording to prevent cross-participant leakage, retaining the top 10 unique ranked chunks based on cosine similarity. By exclusively querying audio and video embedding spaces utilizing the 27 standardized natural-language phrases, we evaluated true cross-modal alignment and explicitly prevented text-to-text lexical retrieval.

**Embedding-model implementation**

We deployed LanguageBind (Zhu et al. 2023), Wave7B (Tang et al. 2025), and Jina Embeddings v5 Omni (Hönicke et al. 2026) as independent inference services accessed through a shared gateway. Every model evaluated identical temporal segments and query phrases zero-shot (Section 2.5). We executed all inference on an NVIDIA RTX 6000 Ada Generation GPU equipped with 48 GB of memory running PyTorch 2.7.1 and CUDA 12.8.

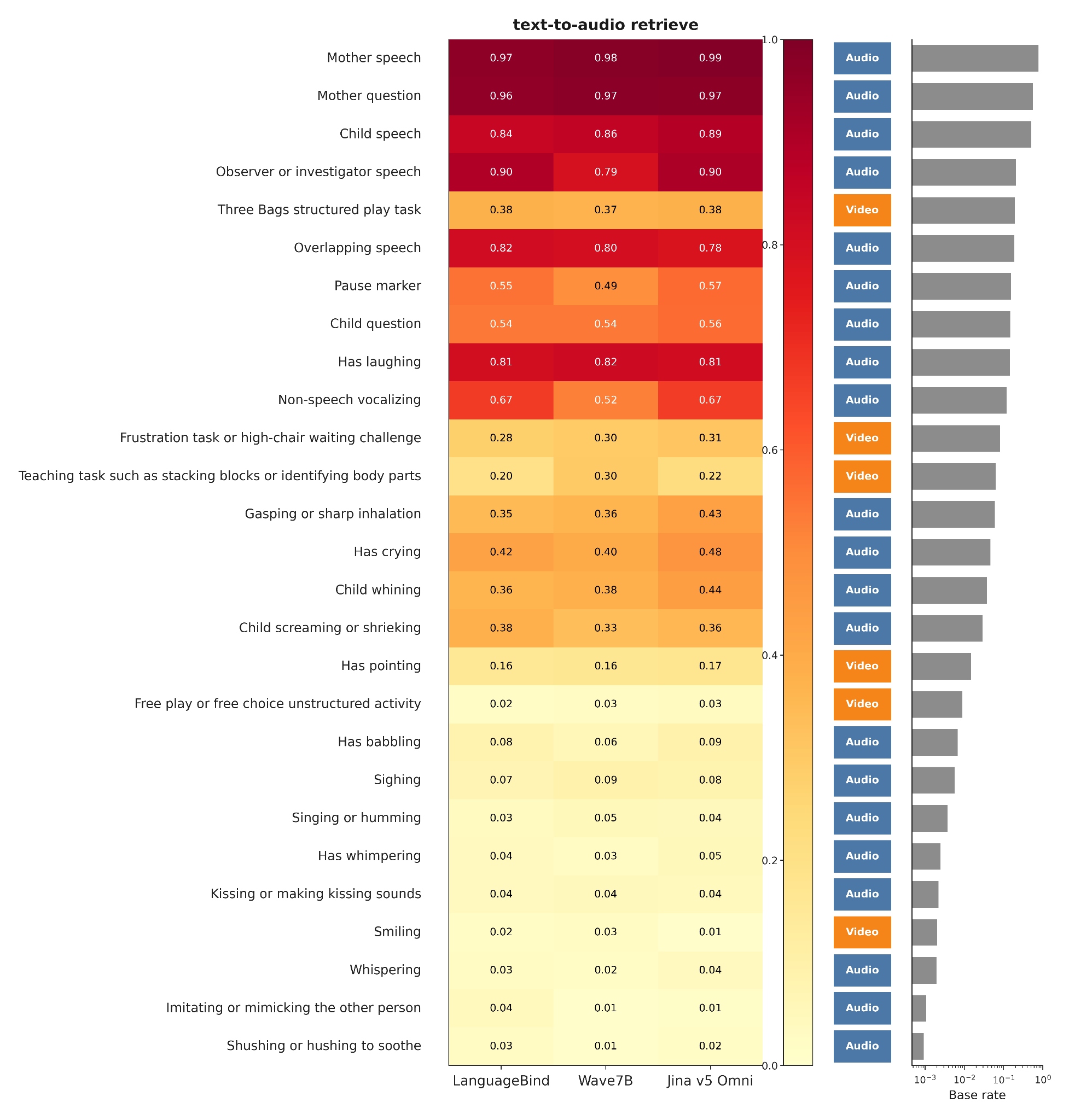

**Supplementary Figure S1.** Per-flag Hit@10 heatmap (text-to-audio retrieve). Rows are the 27 AudioPrimary/VideoPrimary phrase targets sorted by chunk prevalence (highest at top). Columns are embedding models. Side strips mark target taxonomy (Audio vs Video) and log prevalence. Cell values are query-weighted Hit@10. This is the detailed per-flag view summarised in main Figure 4; the companion panel shows the other retrieve channel

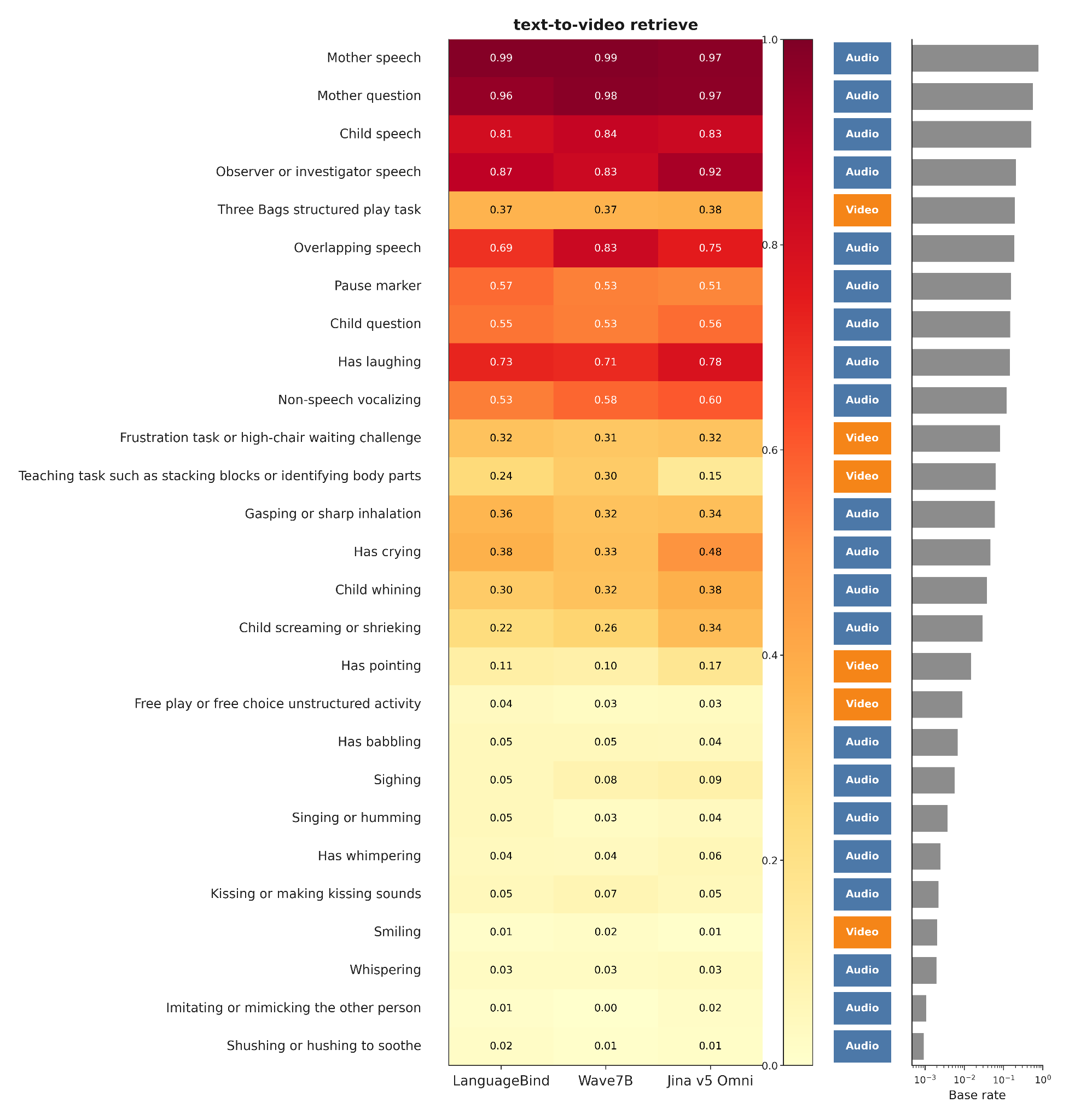

**Supplementary Figure S2.** Per-flag Hit@10 heatmap (text-to-video retrieve). Rows are the 27 AudioPrimary/VideoPrimary phrase targets sorted by chunk prevalence (highest at top). Columns are embedding models. Side strips mark target taxonomy (Audio vs Video) and log prevalence. Cell values are query-weighted Hit@10. This is the detailed per-flag view summarised in main Figure 4; the companion panel shows the other retrieve channel

| **Detail** | **LanguageBind** | **Wave7B** | **Jina v5 Omni** |
| --- | --- | --- | --- |
| Huggingface checkpoint | LanguageBind/LanguageBind_Video_FT,LanguageBind/LanguageBind_Audio_FT | tsinghua-ee/WAVE-7B | jinaai/jina-embeddings-v5-omni-small |
| Architecture class | CLIP-style multimodal contrastive binding | Omni-modal transformer with prediction-embedding extraction | Frozen multimodal tower composition with retrieval adapter |
| Embedding dimension | 768 | 3584 | 1024 |
| Text encoder | CLIP text transformer | Qwen2.5 language model (thinker) | Jina Embeddings v5 text tower |
| Audio encoder | LanguageBind audio encoder | Qwen2.5-Omni audio (Whisper-style) | Frozen Qwen2.5-Omni audio tower |
| Video encoder | LanguageBind video encoder | Integrated vision encoder | Frozen vision tower (Qwen3.5 for small) |
| Embedding extraction | Encoder output | Prediction-embedding layer | Last-token pooling followed by L2 normalization |
| Serving framework | Independent FastAPI service | Independent FastAPI service | FastAPI service using sentence-transformers |
| Fine-tuned on EHS dataset | None | None | None |
| Inference hardware | NVIDIA RTX 6000 Ada, 48 GB | NVIDIA RTX 6000 Ada, 48 GB | NVIDIA RTX 6000 Ada, 48 GB |
| Software environment | PyTorch 2.7.1; CUDA 12.8 | PyTorch 2.7.1; CUDA 12.8 | PyTorch 2.7.1; CUDA 12.8 |

**Supplementary Table S3. Embedding-model implementation and inference environment**. All three models were accessed through the same gateway and evaluated with identical recording segments, queries, and reference labels. All open-weight model checkpoints are available on huggingface.

**Per-chunk metadata and language statistics**

Our preprocessing pipeline preserved detailed chunk-level metadata to support deterministic reference-label generation and language metric reconstruction (Supplementary Table S4). We computed session-level vocabulary diversity strictly from the union of word types across the included transcript windows, deliberately avoiding the sum of chunk-level vocabulary counts to prevent duplicate word scoring.

| **Domain** | **Field** | **Definition** |
| --- | --- | --- |
| Language productivity | Child and maternal NDW | Number of unique lowercased word types within the chunk |
| Language productivity | Child and maternal NTW | Total cleaned word-token count |
| Language productivity | Child and maternal MLU in words | Mean cleaned word tokens per non-zero verbal utterance |
| Language productivity | Child and maternal type-token ratio | NDW divided by NTW |
| Language productivity | Child single-word proportion | Proportion of verbal child utterances containing exactly one word |
| Language productivity | Child and maternal question count | Number of question-terminated utterances |
| Language productivity | Maternal wh-question count | Number of maternal wh-questions |
| Behavioral event | Crying | Counts from &=cries and &=criesandsquirms |
| Behavioral event | Laughing | Counts from &=laughs |
| Behavioral event | Pointing | Counts from &=points and relevant pointing postcodes |
| Behavioral event | Babbling | Counts of phonological fragment tokens and babble markers |
| Behavioral event | Whimpering | Counts from &=whimpers |
| Behavioral event | Vocalizing | Counts from &=sounds, &=grunt, &=noise, and related markers |
| Metadata | Speaker code | CHAT participant code such as CHI, MOT, OBS, or FAT |
| Metadata | Onset and offset | Timing relative to recording onset in milliseconds |
| Metadata | Terminator type | Declarative, question, exclamation, incomplete, trailing, or interrupted |
| Metadata | Activity label | Structured task derived from the active @Activities annotation |
| Metadata | CHAT postcodes | PI, IMP, RD, and other bracketed codes |
| Metadata | Event markers | Dictionary of inline &= event markers and counts |

**Supplementary Table S4. Per-chunk statistics and metadata schema**. The complete-session language reference used all eligible chunks from the recording. Retrieval-derived measures used the same aggregation rules after restricting the calculation to the first 10 unique chunks returned for the child-speech query.

**Retrieval endpoints and cross-model inference**

We calculated all retrieval metrics (Hit@10, Precision@10, Recall@10, MRR, and nDCG@10) and conducted all

statistical inferences (Wilcoxon signed-rank tests and Benjamini–Hochberg / Benjamini–Yekutieli adjustments)

exactly as detailed in the main text (Sections 2.6 and 2.8).

| **Retrieve channel** | **Modality** | **Comparison** | **Metric** | **Participant mean, first model** | **Participant mean, second model** | **Mean paired difference, Δ** | **95% CI for Δ** | **Non-zero pairs, n** | **Rank-biserial r** | **P_BH** | **P_BY** |
| --- | --- | --- | --- | --- | --- | --- | --- | --- | --- | --- | --- |
| text-to-audio | PHRASE_T_A | Jina v5 Omni versus LanguageBind | Hit@10 | 0.3807 | 0.3667 | 0.0140 | [0.0068, 0.0213] | 98 | 0.423 | < 0.001 | 0.001 |
| text-to-audio | PHRASE_T_A | Jina v5 Omni versus Wave7B | Hit@10 | 0.3807 | 0.3589 | 0.0218 | [0.0118, 0.0323] | 105 | 0.443 | < 0.001 | < 0.001 |
| text-to-audio | PHRASE_T_A | Wave7B versus LanguageBind | Hit@10 | 0.3589 | 0.3667 | -0.0078 | [-0.0183, 0.0022] | 105 | -0.098 | 0.407 | 1.000 |
| text-to-audio | PHRASE_T_A | Jina v5 Omni versus LanguageBind | Precision@10 | 0.1576 | 0.1427 | 0.0149 | [0.0123, 0.0175] | 119 | 0.861 | < 0.001 | < 0.001 |
| text-to-audio | PHRASE_T_A | Jina v5 Omni versus Wave7B | Precision@10 | 0.1576 | 0.1456 | 0.0120 | [0.0081, 0.0155] | 119 | 0.623 | < 0.001 | < 0.001 |
| text-to-audio | PHRASE_T_A | Wave7B versus LanguageBind | Precision@10 | 0.1456 | 0.1427 | 0.0029 | [-0.0006, 0.0064] | 116 | 0.192 | 0.098 | 0.326 |
| text-to-audio | PHRASE_T_A | Jina v5 Omni versus LanguageBind | Recall@10 | 0.1129 | 0.0954 | 0.0176 | [0.0132, 0.0215] | 121 | 0.710 | < 0.001 | < 0.001 |
| text-to-audio | PHRASE_T_A | Jina v5 Omni versus Wave7B | Recall@10 | 0.1129 | 0.0953 | 0.0176 | [0.0114, 0.0237] | 121 | 0.551 | < 0.001 | < 0.001 |
| text-to-audio | PHRASE_T_A | Wave7B versus LanguageBind | Recall@10 | 0.0953 | 0.0954 | 0.0000 | [-0.0063, 0.0064] | 121 | 0.101 | 0.384 | 1.000 |
| text-to-audio | PHRASE_T_A | Jina v5 Omni versus LanguageBind | MRR | 0.2355 | 0.2094 | 0.0262 | [0.0184, 0.0334] | 121 | 0.657 | < 0.001 | < 0.001 |
| text-to-audio | PHRASE_T_A | Jina v5 Omni versus Wave7B | MRR | 0.2355 | 0.2071 | 0.0284 | [0.0206, 0.0362] | 121 | 0.644 | < 0.001 | < 0.001 |
| text-to-audio | PHRASE_T_A | Wave7B versus LanguageBind | MRR | 0.2071 | 0.2094 | -0.0022 | [-0.0102, 0.0061] | 121 | -0.047 | 0.655 | 1.000 |
| text-to-audio | PHRASE_T_A | Jina v5 Omni versus LanguageBind | nDCG@10 | 0.1830 | 0.1604 | 0.0226 | [0.0182, 0.0268] | 121 | 0.853 | < 0.001 | < 0.001 |
| text-to-audio | PHRASE_T_A | Jina v5 Omni versus Wave7B | nDCG@10 | 0.1830 | 0.1627 | 0.0203 | [0.0143, 0.0262] | 121 | 0.641 | < 0.001 | < 0.001 |
| text-to-audio | PHRASE_T_A | Wave7B versus LanguageBind | nDCG@10 | 0.1627 | 0.1604 | 0.0024 | [-0.0031, 0.0078] | 121 | 0.178 | 0.112 | 0.371 |
| text-to-video | PHRASE_T_V | Jina v5 Omni versus LanguageBind | Hit@10 | 0.3596 | 0.3424 | 0.0172 | [0.0079, 0.0266] | 97 | 0.442 | < 0.001 | < 0.001 |
| text-to-video | PHRASE_T_V | Jina v5 Omni versus Wave7B | Hit@10 | 0.3596 | 0.3479 | 0.0117 | [0.0018, 0.0211] | 106 | 0.303 | 0.008 | 0.027 |
| text-to-video | PHRASE_T_V | Wave7B versus LanguageBind | Hit@10 | 0.3479 | 0.3424 | 0.0055 | [-0.0049, 0.0166] | 101 | 0.112 | 0.348 | 1.000 |
| text-to-video | PHRASE_T_V | Jina v5 Omni versus LanguageBind | Precision@10 | 0.1587 | 0.1397 | 0.0190 | [0.0156, 0.0222] | 120 | 0.849 | < 0.001 | < 0.001 |
| text-to-video | PHRASE_T_V | Jina v5 Omni versus Wave7B | Precision@10 | 0.1587 | 0.1469 | 0.0118 | [0.0076, 0.0161] | 121 | 0.544 | < 0.001 | < 0.001 |
| text-to-video | PHRASE_T_V | Wave7B versus LanguageBind | Precision@10 | 0.1469 | 0.1397 | 0.0073 | [0.0027, 0.0118] | 119 | 0.370 | < 0.001 | 0.002 |
| text-to-video | PHRASE_T_V | Jina v5 Omni versus LanguageBind | Recall@10 | 0.1072 | 0.0848 | 0.0224 | [0.0169, 0.0275] | 121 | 0.738 | < 0.001 | < 0.001 |
| text-to-video | PHRASE_T_V | Jina v5 Omni versus Wave7B | Recall@10 | 0.1072 | 0.0871 | 0.0201 | [0.0136, 0.0264] | 121 | 0.646 | < 0.001 | < 0.001 |
| text-to-video | PHRASE_T_V | Wave7B versus LanguageBind | Recall@10 | 0.0871 | 0.0848 | 0.0023 | [-0.0034, 0.0086] | 121 | 0.064 | 0.544 | 1.000 |
| text-to-video | PHRASE_T_V | Jina v5 Omni versus LanguageBind | MRR | 0.2247 | 0.2000 | 0.0247 | [0.0168, 0.0327] | 121 | 0.603 | < 0.001 | < 0.001 |
| text-to-video | PHRASE_T_V | Jina v5 Omni versus Wave7B | MRR | 0.2247 | 0.2083 | 0.0164 | [0.0074, 0.0253] | 121 | 0.432 | < 0.001 | < 0.001 |
| text-to-video | PHRASE_T_V | Wave7B versus LanguageBind | MRR | 0.2083 | 0.2000 | 0.0083 | [-0.0004, 0.0174] | 121 | 0.184 | 0.091 | 0.302 |
| text-to-video | PHRASE_T_V | Jina v5 Omni versus LanguageBind | nDCG@10 | 0.1800 | 0.1532 | 0.0269 | [0.0217, 0.0322] | 121 | 0.819 | < 0.001 | < 0.001 |
| text-to-video | PHRASE_T_V | Jina v5 Omni versus Wave7B | nDCG@10 | 0.1800 | 0.1621 | 0.0179 | [0.0112, 0.0243] | 121 | 0.581 | < 0.001 | < 0.001 |
| text-to-video | PHRASE_T_V | Wave7B versus LanguageBind | nDCG@10 | 0.1621 | 0.1532 | 0.0089 | [0.0027, 0.0156] | 121 | 0.302 | 0.005 | 0.018 |

**Supplementary Table S5. Participant-level pairwise comparisons of retrieval performance in the combined sample (dual retrieve channels)** Paired Wilcoxon signed-rank tests on participant-level metric means (n = 121 participants). Each block is one retrieve channel (text-to-audio / PHRASE_T_A or text-to-video / PHRASE_T_V); within a channel, each row is one model contrast within one of five principal retrieval measures (Hit@10, Precision@10, Recall@10, MRR, nDCG@10). Positive Δ and rank-biserial r favour the first-named model. P_BH and P_BY are Benjamini–Hochberg and Benjamini–Yekutieli adjustments across the 15 aggregate tests within each channel (as written in the source pairwise CSVs). Values of P < 0.001 are shown as “< 0.001”. Main-text Table 3 reports the Hit@10 subset. Query-weighted means in Figure 2 (text-to-audio: n = 7479; text-to-video: n = 7479) are a separate descriptive summary and are not reported in this table.

| **Specification** | **Estimand** | **n** | **β / month** | **95% CI** | **Mapped Δ (14→36)** | **P** | **Interpretation** |
| --- | --- | --- | --- | --- | --- | --- | --- |
| Primary (all-available) | Target-present | 94 | 0.00299 | [0.00151, 0.00446] | +0.066 | < 0.001 | Age-related gain |
| Complete-case sensitivity | Target-present | 62 | 0.00303 | [0.00132, 0.00474] | +0.067 | < 0.001 | Age-related gain |
| Secondary (all-query) | All queries | 94 | -0.00197 | [-0.00294, -0.00100] | -0.043 | < 0.001 | Apparent decline (prevalence-confounded) |
| Speech ablation | Target-present; excl. child & maternal speech | 94 | 0.00175 | [0.00012, 0.00338] | +0.039 | 0.035 | Attenuated gain |
| Infant-paralinguistic ablation (n = 8) | Target-present; excl. 8 early vocal/affective targets | 94 | 0.00525 | [0.00382, 0.00668] | +0.116 | < 0.001 | Steepened gain |

**Supplementary Table S6. Jina Hit@10 longitudinal GEE sensitivity (dual retrieve channels pooled)**. Continuous-age GEE coefficients for Jina Embeddings v5 Omni Hit@10 (Gaussian family, identity link, exchangeable working correlation, robust SEs, participant clustering). Primary and ablation rows use target-present queries among participants assessed at two or more waves (n = 94). Mapped Δ = β × 22 months (14→36 mo). The secondary all-query row restores target-absent queries (Hit@10 = 0 by definition when the behaviour is absent). Speech ablation excludes child and maternal speech; infant-paralinguistic ablation excludes babbling, crying, gasping, laughing, screaming, non-speech vocalizing, whimpering, and whining. Model × time interaction (joint Wald on the three-model target-present GEE): χ²(4) = 10.00, P = 0.040 (n_clusters = 94). These specifications support main-text Section 3.4 and Figure 5.

### Within-wave comparison of retrieval-derived and complete-session language measures

As detailed in the main text (Sections 2.7 and 2.8), we calculated retrieval-derived and complete-session language metrics utilizing identical measurement rules.

Crucially, we aggregated child vocabulary diversity (CVD) by taking the union of distinct lowercased word types across the included utterances, explicitly

avoiding the summation of chunk-level vocabulary counts to prevent duplicate word scoring. We aggregated mean length of utterance (MLU) and single-word

proportion (SWP) utilizing the number of verbal child utterances as weights, rendering them undefined when retrieved chunks contained no eligible verbal

utterances. To prevent expected age-related language increases from confounding correlations, we conducted all comparisons strictly within distinct developmental waves. Primary language validation used text-to-audio retrieval of child-speech queries for Jina Embeddings v5 Omni and did not pool text-to-video rankings (main-text Section 3.5). Within each wave, recordings lacking eligible verbal child utterances in the retrieved subset were excluded from MLU and SWP (at 14 months: CVD n = 105; MLU/SWP n = 75). We applied Spearman correlation to quantify rank-order preservation, adding Pearson correlation as a secondary measure of linear correspondence. We utilized mean absolute error (MAE) to capture average difference magnitudes and signed error to detect systematic biases, where negative values explicitly indicate underestimation of the complete-session measure. Finally, we estimated confidence intervals for Spearman correlations utilizing 500 recording-level bootstrap resamples within each wave.
